# Resting State Functional Connectivity Demonstrates Increased Segregation in Bilateral Temporal Lobe Epilepsy

**DOI:** 10.1101/2022.08.11.22278372

**Authors:** Alfredo Lucas, Eli J. Cornblath, Nishant Sinha, Peter Hadar, Lorenzo Caciagli, Simon S. Keller, Leonardo Bonilha, Russell T. Shinohara, Joel M. Stein, Sandhitsu Das, Ezequiel Gleichgerrcht, Kathryn A. Davis

## Abstract

Temporal lobe epilepsy (TLE) is the most common type of focal epilepsy. An increasingly identified subset of patients with TLE consists of those who show bilaterally independent temporal lobe involvement during seizures. Bilateral TLE (BiTLE) remains understudied, likely due to its complex underlying pathophysiology and heterogeneous clinical presentation.

In this study, using a multicenter resting state functional MRI (rs-fMRI) dataset, we constructed whole brain functional networks of 19 patients with BiTLE, and compared them to those of 75 patients with unilateral TLE (UTLE). We quantified resting-state, whole-brain topological properties using metrics derived from network theory, including clustering coefficient, global efficiency, participation coefficient, and modularity. For each metric, we computed an average across all brain regions, and iterated this process across network densities ranging from 0.10-0.50. Curves of network density versus each network metric were compared between groups. Finally, we derived a combined metric, which we term the “integration-segregation axis”, by combining whole brain average clustering coefficient and global efficiency curves and applying principal component analysis (PCA)-based dimensionality reduction.

Compared to UTLE, BiTLE had decreased global efficiency (p=0.026), increased whole brain average clustering coefficient (p=0.035), and decreased whole brain average participation coefficient across a range of network densities (p=0.001). Modularity maximization yielded a larger number of smaller communities in BiTLE than in UTLE (p=0.016). Differences in network properties separate BiTLE and UTLE along the integration-segregation axis: 68% of patients with BiTLE were identified within the high segregation region, while only 41% of the UTLE patients were identified in the same region (p=0.042). Along the integration-segregation axis, UTLE patients with poor surgical outcomes were more similar to BiTLE than those with good surgical outcomes (p=0.72).

Increased interictal whole brain network segregation, as measured by rs-fMRI, is specific to BiTLE, and may assist in non-invasively identifying this patient population prior to intracranial EEG or device implantation.

## Introduction

The most common subtype of focal epilepsy is temporal lobe epilepsy (TLE)^1^, which is characterized by seizures originating from the medial and/or lateral regions of the temporal lobes. For patients who continue to have seizures despite medical management, tailored resection of brain structures involved in TLE is associated with overall good post-surgical prognosis^2,3^. However, one subset of TLE patients poses a particularly challenging clinical entity: those who show bilaterally independent temporal lobe seizure onset.

Bilateral temporal lobe epilepsy (BiTLE) remains an understudied and complex clinical entity. Correct identification of BiTLE in patients without radiographic signs of underlying bilateral mesial temporal sclerosis (MTS), is generally possible with intracranial electroencephalography (iEEG)^4^. In a recent study, up to 70% of patients thought to have BiTLE based on scalp EEG were then proven to have unilateral TLE (UTLE) after iEEG^5^. Other authors have explored the anatomical and electroclinical phenotypes of BiTLE^5–8^, as well as related presurgical evaluation and surgical strategies^5,9,10^. Despite the available literature, there is still no consensus regarding the appropriate diagnosis and best therapeutic intervention for BiTLE. Finally, there is a gap in the current literature regarding the underlying brain network structure that characterizes BiTLE, or how this network differs between BiTLE and UTLE.

Epilepsy has been increasingly identified as a network disorder^11–13^. The study of brain connectomics has demonstrated the presence of large-scale network abnormalities even in focal epilepsies^14–16^. Widespread epileptic networks can be observed clinically, particularly in cases where surgical resection of the suspected seizure foci does not lead to post-surgical seizure freedom^17^. Non-invasive imaging modalities, such as resting state functional MRI (rs-fMRI), have revealed both focal and global epileptic network abnormalities^18–20^. Further study of these network abnormalities may provide biomarkers for the classification and localization of epilepsy subtypes, and may help to guide therapeutic decision making noninvasively^21^.

In addition to their promise as biomarkers, network metrics are well poised for characterizing both short- and long-range interactions of brain regions. Quantifying network integration, that refers to how interconnected different parts of the brain are, as well as network segregation, that refers to how independent different parts of the brain are, is tractable in network neuroscience. The balance between segregation and integration of brain networks has been studied in the setting of normal brain activity^22–25^, aging^26^, Alzheimer’s disease^27^, and several neuropsychiatric disorders^28,29^. In epilepsy, both animal^30^ and human studies^19,31^ have shown increased segregation of epileptic networks relative to controls. Nevertheless, few studies have focused on the integration-segregation balance in patients within different epilepsy subtypes. To our knowledge, this question remains unexplored in BiTLE. Specifically, it remains unclear whether the independent seizure onset from both hemispheres is a consequence of multiple epileptogenic subnetworks acting independently, or a larger epileptogenic network with widespread involvement of both hemispheres. In other words --are the networks in BiTLE highly integrated, or highly segregated?

In this study, we aim to characterize the whole-brain topological properties of interictal resting state networks in BiTLE using resting-state functional MRI (rs-fMRI). We hypothesize that BiTLE and UTLE have different functional network integration and segregation profiles, and that these differences can be quantified through network-based statistics. We further hypothesize that these differences in network topology can distinguish these two populations of TLE patients in a data-driven manner. Our findings suggest that BiTLE has higher whole brain network segregation when compared to UTLE. We also find that UTLE patients with poor surgical outcomes have similar integration-segregation profiles as BiTLE patients when compared to those with good surgical outcomes. Our results shed light on the underlying network architecture of BiTLE relative to UTLE and provide a network-centric view of BiTLE that may enable stratification of patients during presurgical evaluation.

## Methods

### Subject Demographics

Data acquisition for this study was approved by the institutional review boards of the University of Pennsylvania (Penn) and the University of Liverpool (UOL). We compiled a multi-center dataset consisting of 94 drug-resistant TLE patients who had undergone neuroimaging during presurgical evaluation including resting state functional magnetic resonance imaging (fMRI).

From Penn, we studied 77 patients. Localization of seizure focus was determined during the Penn Epilepsy Surgical Conference (PESC Lateralization and PESC Localization) following evaluation of various clinical, neuroimaging, and neurophysiological data including: seizure semiology, neuropsychological testing, MRI, positron emission tomography (PET), magnetoencephalography (MEG), scalp EEG, and intracranial EEG findings. The PESC is a multi-disciplinary team that consists of neurologists, neurosurgeons, neuroradiologists, neuropsychologists, and nuclear medicine specialists. Thirty-four patients had left-sided lateralization, 24 patients had right-sided lateralization, and 19 patients were deemed to have bilaterally independent onset TLE. Age, gender, MRI lesional status, lateralization and Engel surgical outcome data^32^, where available, are reported in **Table 1**. Engel scores of IA-D were considered good seizure outcomes and scores of II-IV were considered poor seizure outcomes. The MRI, EEG and PESC results, as well as the detailed surgical outcomes, are provided in the **Supplementary Table 1**. All subject identifiers included in **Supplementary Table 1** were randomly generated and were only known to the research group.

**Table 1:**
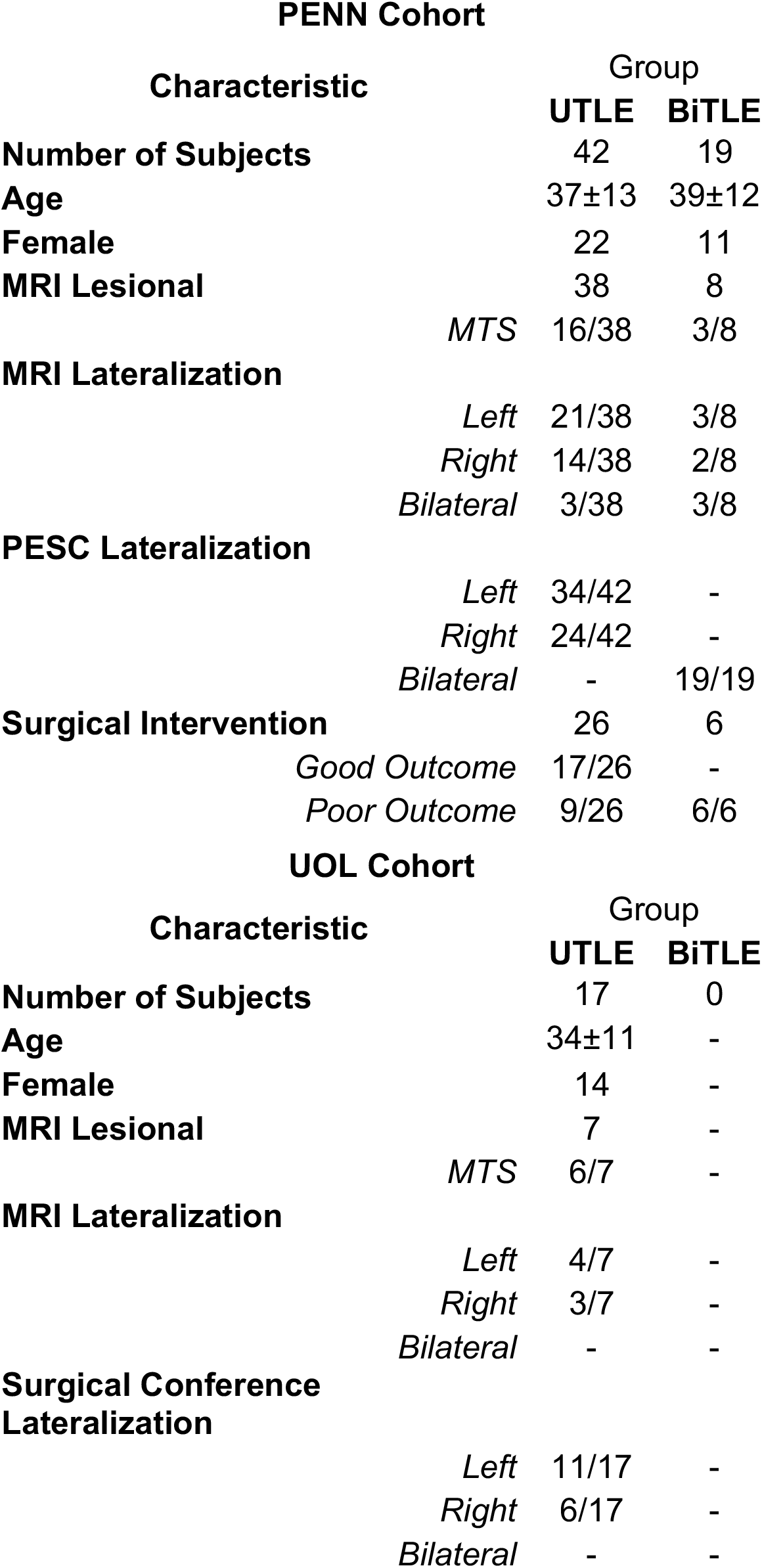
Summary characteristics for unilateral temporal lobe epilepsy (UTLE) and bilateral temporal lobe epilepsy (BiTLE) subjects included in this study. MTS: mesial temporal sclerosis. PESC: Penn Epilepsy Surgical Conference.

An additional cohort of 17 drug resistant TLE patients were included from UOL. Localization and seizure focus was determined through a multidisciplinary surgical conference as with the Penn Cohort. All the patients in this cohort were unilateral TLE patients with 11 left-sided, and 6 right-sided seizure onset (SOZ) lateralization. Age, gender, MRI lesional status, and final lateralization are summarized in **Table 1**.

### Image Acquisition

For 63/77 patients of the Penn cohort, we used a Siemens 3T Magnetom PrismaFit scanner. Resting-state fMRI data were acquired during a 9-min interval with an axial, 72-slice gradient echo-planar sequence, TE/TR=37**/**800ms, with a 2mm isotropic voxel size (Penn protocol 1). For the remaining 14/77 patients of the Penn cohort, we used a Siemens 3T Magnetom Trio scanner. For this subset of patients, resting-state fMRI data were acquired during a 6-min interval with an axial, 72-slice gradient echo-planar sequence, TE/TR=37**/**800ms, with a 2mm isotropic voxel size (Penn protocol 2). High-resolution T1-weighted images, with a sagittal, 208-slice MPRAGE sequence, TE/TR=2.24/2400ms, with a 0.8mm isotropic voxel size were acquired in all participants.

For all participants from the UOL cohort, we used a 3T GE Discovery 750 scanner. T1-weighted data were acquired using the following parameters: Pulse sequence = BRAVO; echo time (TE) = 3.22 ms; repetition time (TR) = 8.2 ms; field of view (FOV) = 24, TI = 450 ms; slice thickness = 1 mm; voxel size = 1 mm × 1 mm × 1 mm; 140 slices; flip angle = 12. rs-fMRI data were acquired during a 6-min interval with a T2-weighted sequence with the following parameters: Pulse sequence = gradient echo; TE = 25 ms; TR = 2000 ms; FOV = 24; slice thickness = 2.4 mm; voxel size = 3 mm × 3 mm × 3 mm; 180 volumes; 38 slices; flip angle = 75.

At both sites, participants were instructed to stay awake and to look at a white fixation cross on a black background.

### Neuroimaging Processing

We used fMRIPrep^33^ to perform brain extraction and segmentation of individual T1-weighted (T1w) images, registration of rs-fMRI data to individual T1w and MNI template space, and time-series confound estimation. We included complete details about the functional and anatomical processing pipelines implemented by fMRIPrep in the **Supplementary Methods**. We used the fMRIPrep output data as our input to the xcpEngine post-processing pipeline for confound regression, demeaning, detrending and temporal filtering^34^. For our regression parameters, we included motion realignment parameters (3 rotational and 3 translational), the mean white matter and cerebrospinal fluid time series over all voxels^35^, the mean time series signal across the whole brain (global signal regression)^36^, the temporal derivative of motion parameters^35^, and finally, the second power of each of the previously mentioned regressors (a total of 36 regression parameters). The timeseries and regressors were detrended and band-passed between 0.01-0.10Hz. The filtered regressors were fitted to the filtered BOLD timeseries data using multiple linear regression.

### Functional Connectivity

A complete overview of our methodological pipeline is shown in **Figure 1**. We calculated the average of BOLD timeseries for each voxel within each parcel of the Brainnetome atlas (210 cortical and 36 subcortical parcels)^37^. From the mean timeseries at each parcel, we created a 246×246 functional connectivity matrix by taking the Pearson correlation between timeseries. For the UTLE patients, we left-right flipped the functional connectivity matrices such that the regions ipsilateral to the SOZ would match across subjects. Since our dataset consisted of patients from two different institutions with different fMRI acquisition parameters, we harmonized the final functional connectivity matrices for the UOL cohort and the Penn cohort scanned under Penn protocol 2 to the Penn protocol 1 using NeuroCombat^38^. We also included the group assignments (UTLE and BiTLE) as a covariate to preserve in the NeuroCombat model. During harmonization each connection of the functional connectivity matrix was harmonized separately across subjects.

**Figure 1.**
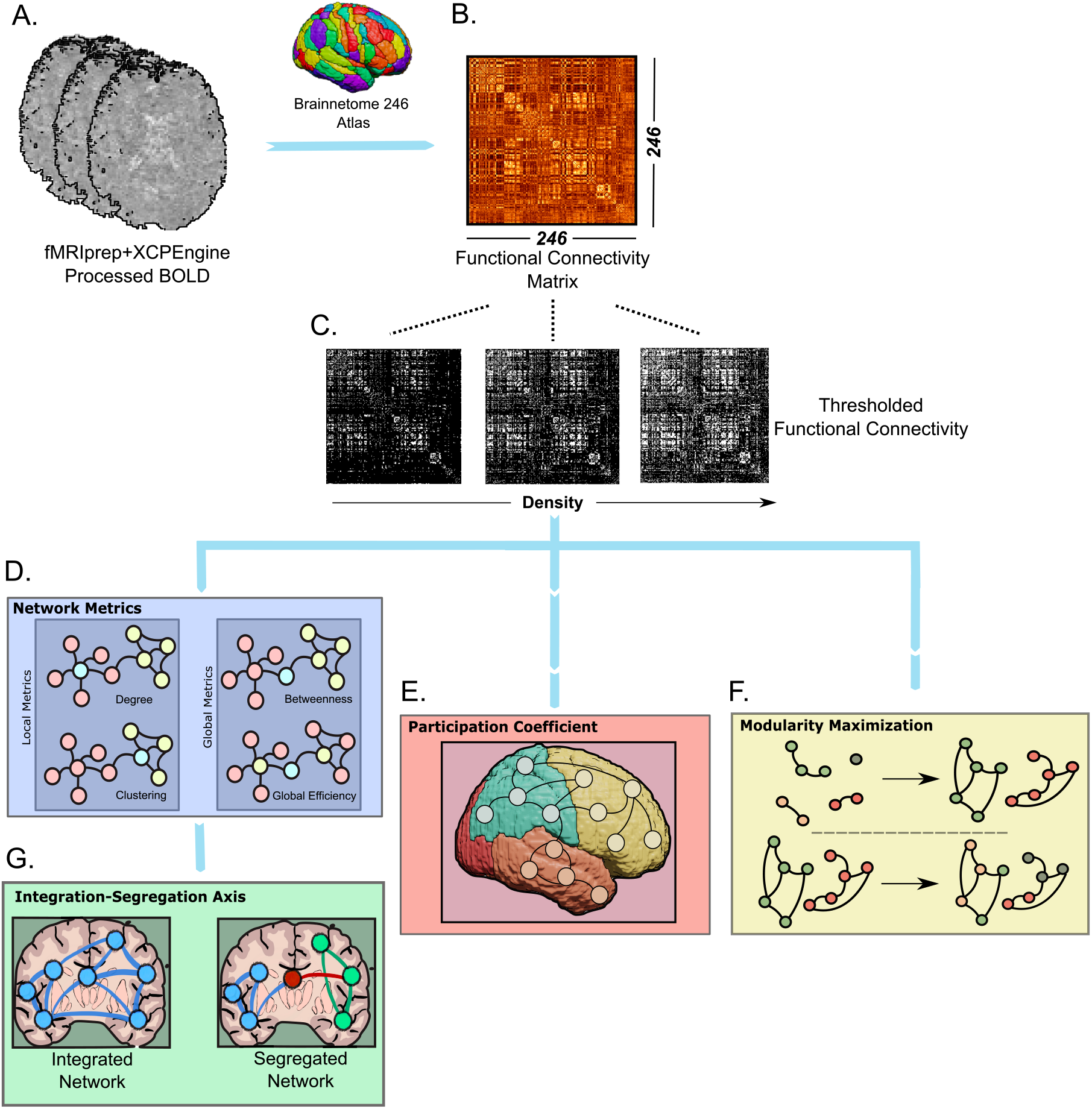
Methods Outline: A. fMRI data was processed with fMRIprep+XCPEngine and subsequently parcellated into the Brainetomme atlas. B. For each subject, a functional connectome was constructed via Pearson correlations between ROIs defined by the atlas. C. Each functional connectome was subsequently thresholded into network densities ranging from 0.10-0.50. D. The thresholded connectomes were used to compute whole brain average degree, clustering, betweenness, as well as global efficiency, generating density dependent curves for each of these metrics. The participation coefficient I and the number of communities detected through modularity maximization (F) were also estimated from the thresholded connectomes in a density-dependent manner. Finally, using the whole brain average clustering and global efficiency density curves an integration-segregation axis was generated (G).

### Network Metrics

A common approach to assess the topological properties of functional connectomes is to threshold the weights of the edges to minimize the presence of spurious connections. However, there is no consensus on the optimal threshold and the resulting binarized matrices can be biased depending on the chosen threshold value. To address this caveat and avoid the use of an arbitrary threshold^19,39–42^, we conducted our analyses in connection-density-dependent manner. Specifically, network topological properties were estimated at proportional thresholds representing connection densities ranging from 10% to 50%, with 1% increments^43^. During thresholding, the absolute value of all connections was taken. We chose the lower bound of 10% since networks tend to become disconnected at low densities, and the upper bound of 50% since network properties at higher densities tend to randomness due to the inclusion of potentially confounding associations^39,40^. By implementing this approach, we obtained 40 values per each metric, per patient.

At each network density, we computed the following metrics: degree, betweenness centrality, clustering coefficient and global efficiency^44,45^. These network metrics, measured through rs-fMRI, have been demonstrated to vary depending on patients’ epilepsy subtype^20,46,47^ and lateralization of the SOZ^48,49^. Nodal metrics, namely the degree, betweenness centrality, and clustering coefficient, were computed at each node (or parcellation), and subsequently averaged across all nodes in the network, resulting in a single global average value for each metric across densities. All network metrics, as well as the participation coefficient and modularity calculations, were carried out using a Python 3.8 implementation of the MATLAB Brain Connectivity Toolbox^50^.

### Participation Coefficient

We computed the participation coefficient^51^ to directly assess the degree of integration/segregation of putative functional and anatomic brain communities. Functional community assignments provide a natural framework for studying participation across communities, since these assignments are based on rs-fMRI data^52^. Anatomic community assignments are useful in this setting since localizing network abnormalities within a specific anatomic location (e.g., temporal lobe in TLE) is of particular interest in epilepsy. Each of the 246 parcellations in the Brainnetome atlas was assigned a) a functional community based on the Yeo-Krienen 7 cortical networks^52^, and b) an anatomic community from among frontal, parietal, temporal, insular, occipital, limbic or subcortical localization defined by the lobar assignments of the Brainnetome atlas. Both the functional and anatomic assignments were further split into left/right, for the BiTLE subjects, and ipsilateral/contralateral for the UTLE subjects. Therefore the final analysis looked at both left and right communities for each of the main communities (14 total communities for the functional and 14 total communities for the anatomic community definitions). As with the other network metrics, the participation coefficient was computed at each node and then subsequently averaged either across the entire brain or within each community. This process was repeated across the same range of densities defined previously.

### Modularity Maximization

Modularity maximization was carried out to identify the number of communities that could independently be detected for each participant, with the assumption that a network with a larger number of communities has a higher degree of segregation. The Louvain algorithm for modularity maximization was utilized^53^. Given that the communities detected were different for each patient, the number of communities detected was the metric of interest. The community detection was performed in two ways. First, we performed community detection in a network density-dependent manner, with the commonly used resolution parameter of *γ*=1. Second, we identified communities in the unthresholded functional connectome. This was done at a range of *γ* values between 0.5-3.0 with a spacing of 0.1, as we expected divergence between the two groups to happen in a scale-dependent manner^54^. The first approach quantifies how the number of communities detected in the two groups differ as the size of the network itself gets larger (higher densities, larger network). The second approach quantifies how the number of communities detected in BiTLE and UTLE diverge as the detected communities get smaller (larger γ, smaller communities).

### Integration-Segregation Axis

To characterize the spectrum of integration and segregation in UTLE and BiTLE, we implemented a strategy for generating an integration-segregation axis based on the average clustering coefficient and the global efficiency (**Figure 2)**. For each subject, the average clustering coefficient and global efficiency density curves were concatenated, resulting in a Nx80 dimensional matrix, where N is the number of subjects, and 80 the concatenated dimension (40 values for clustering coefficient across densities and 40 values for global efficiency across densities). Both BiTLE and UTLE patients were used, so the final matrix was 95×80. We performed principal component analysis (PCA), and the first two principal components (PCs) were kept. Only the first two PCs were kept since the variance explained by the first PC was 96%, and the variance explained by the second PC was 1.9%, suggesting that most of the explanatory power was concentrated in the first PC (**Supplementary Figure 1A**).

**Figure 2.**
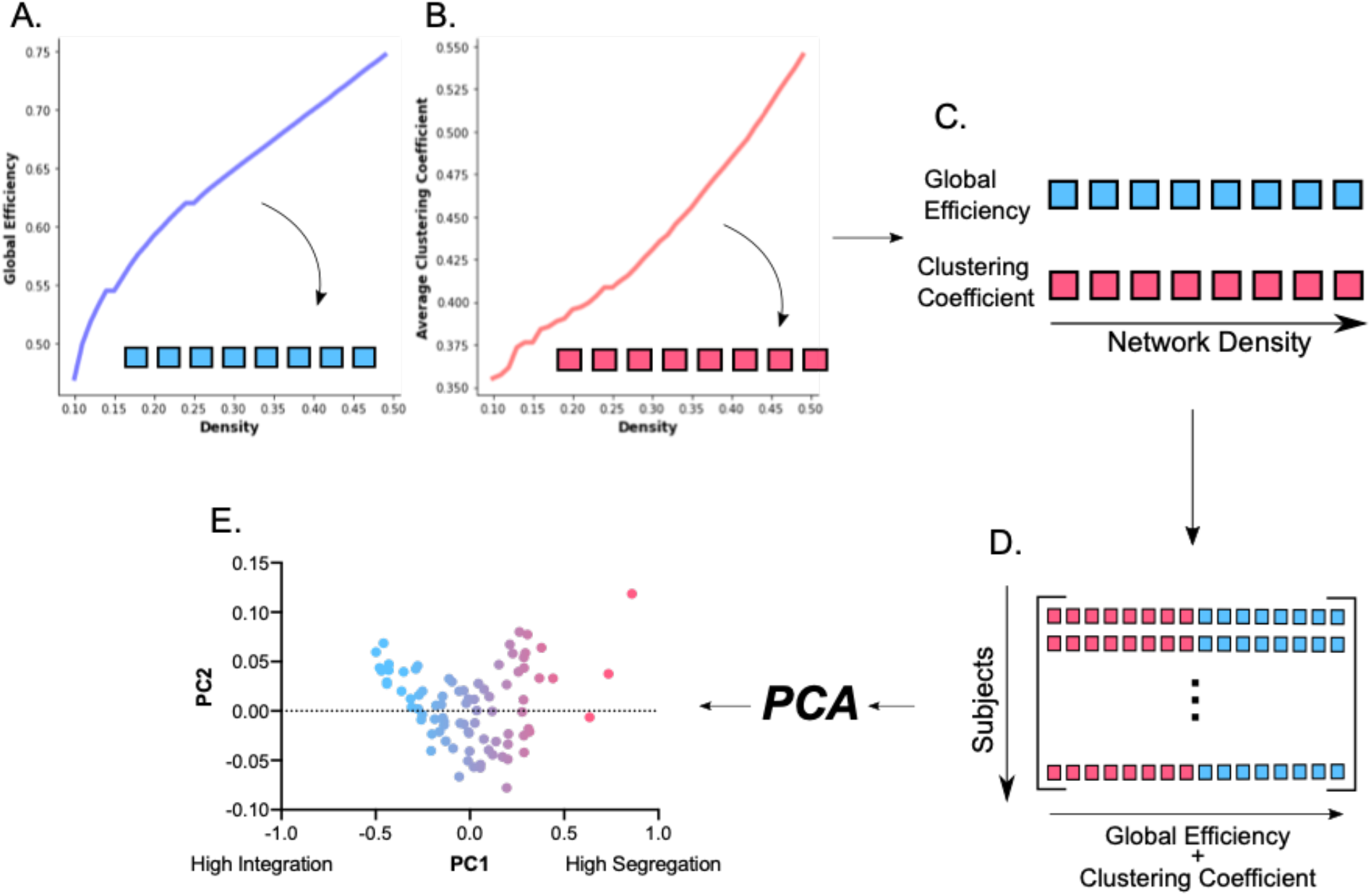
Generation of the integration-segregation axis: For each subject, the global efficiency (A) and average clustering coefficient (B) density curves are vectorized (C). The two vectorized curves are subsequently concatenated and the resulting vector forms the rows of the integration-segregation matrix (D), with one row per subject. After applying PCA to this matrix, we generate the integration-segregation axis by taking the first two PCs (E).

Analysis of the loadings of the first two components showed that the first component was effectively the spectrum of integration and segregation as measured by the tradeoff between global efficiency and average clustering coefficient (**Supplementary Figure 1B**). Given this, negative values of the first principal component suggested high integration, whereas positive values suggested high segregation. The second principal component provided a linear combination across densities between the two metrics, yielding larger values (more positive) for larger densities (**Supplementary Figure 1C**). After generating the integration-segregation axis, spectral clustering was used to identify two clusters, a high-integration and a high-segregation cluster. The cluster assignments for each patient group were quantified, providing an estimate for how many UTLE and BiTLE patients belong to each cluster. Finally, using the same cluster assignments, the number of UTLE patients with good and poor surgical outcomes were quantified in each of the two clusters. Both PCA and spectral clustering were carried out in Sklearn version 1.0.2.

### Statistical Analysis

Statistical comparison of the network density dependent curves of average degree, average clustering, average betweenness, global efficiency, average participation and number of communities detected through modularity maximization was done through a 1-dimensional suprathreshold cluster detection procedure, analogous to the one proposed Nichols et al.^55^. This approach is commonly used in the fMRI literature and provides family wise error rate (FWER) correction without sacrificing too much power. Briefly, for a given network metric at each density, a Welch’s *t*-test was carried out, generating a *t*-statistic at each density. This 1-dimensional vector of t-statistics was then thresholded at a significance level of p<0.05, uncorrected. Densities above the threshold were identified, and 1-dimensional clusters, consisting of groups of adjacent densities that surpassed the threshold, were defined. Then, the group labels for each subject were permuted 1000 times, and the process above was repeated for each permutation, generating 1000 different sets of 1-dimensional clusters. From the permuted clusters, a distribution consisting of the maximum cluster size in each permutation was generated. Cluster sizes from the unpermuted data that were larger than 95% of the maximum permuted cluster sizes were considered significant (equivalent to p<0.05). The p-value of this significant cluster (p_cluster_) is then estimated as the proportion of permuted cluster sizes that are larger than the largest cluster size in the unpermuted data, with the addition of a positive bias^56^. This procedure is well suited for our specific application for the following reasons. First, the differences in fMRI seen in epilepsy have small effect sizes^31^, which requires the use of strategies that can increase statistical power, particularly in relatively small sample sizes. Second, we are interested in answering the question of whether there is a range of network densities in which the measured network properties differ between the groups, and not necessarily at which specific densities this happens. The above clustering procedure increases statistical power, but it does so at the expense of only determining whether a cluster of densities is significant or not, and not which specific density is significant. For each metric, we also measured the absolute value of the Cohen’s D between the two groups at each density to quantify effect sizes. We considered Cohen’s D values above 0.5 to represent a moderate effect size, and above 0.8 to represent a large effect size^57^.

In order to identify which nodes of the network had significant differences between the groups for a given density and network metric, we applied the same permutation-based cluster-wise procedure as above, but across a set of network nodes. This approach is analogous to that used in past studies^39,58^, and a detailed description of our implementation is included in the **Supplementary Methods**.

Results were considered statistically significant for p<0.05. Density-dependent curves are plotted with the mean as a dashed line and the 95% confidence interval shaded in the respective color.

### Data Availability Statement

Fully processed functional connectivity matrices for all the subjects included in the study are available here: https://doi.org/10.26275/t8ls-zuw2. The code used to perform all analyses and generate all figures is available on GitHub at: https://github.com/allucas/BiTLE_Segregation.

## Results

### Network Metrics Show Increased Segregation in BiTLE

We expected differences between the network metrics of UTLE and BiTLE to be present across network densities. To assess this, we quantified whole brain average degree, average betweenness centrality, average clustering coefficient, and global efficiency across network densities between 0.10-0.50 and compared the resulting curves between groups (**Figure 3)**. For both groups, average degree, global efficiency, and average clustering increased as a function of network density, whereas the average betweenness centrality decreased. No significant differences were seen between BiTLE and UTLE for average degree (p_cluster_=0.102). There were differences in average betweenness centrality with a moderate effect size between densities of 0.15-0.30 (**Supplementary Figure 3B**), with statistically significant higher average betweenness for BiTLE (**Figure 3B**) between densities of 0.12-0.18 (p_cluster_=0.041). Differences in global efficiency had moderate effect sizes between densities of 0.10-0.30 (**Supplementary Figure 3C**), with statistically significant lower global efficiency in BiTLE between densities of 0.10-0.17 (p_cluster_=0.026) (**Figure 3C**). Finally, differences in average clustering coefficient had moderate effect sizes between densities of 0.20-0.50 (**Supplementary Figure 3D**), with higher average clustering in BiTLE across that range (**Figure 3D**), and a statistically significant higher average clustering in BiTLE between densities of 0.06-0.32 (p_cluster_=0.035). Overall, our results suggest decreased global efficiency and increased clustering and betweenness across a range of densities for BiTLE when compared to UTLE.

**Figure 3.**
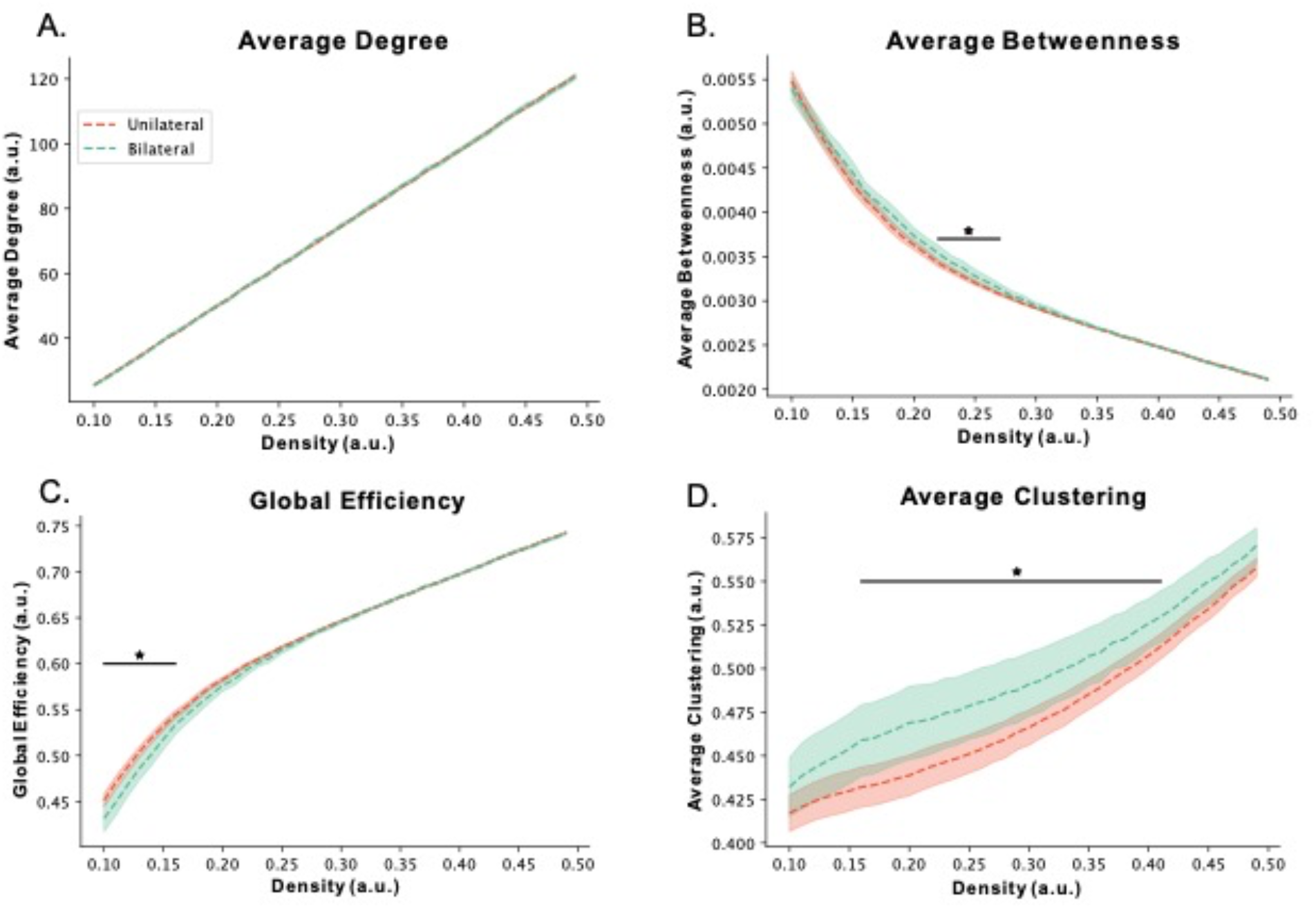
Network Metrics: Whole-brain A. average degree, B. average betweenness (significantly different between densities of 0.12-0.18), C. global efficiency (significantly different between densities of 0.10-0.19) and D. average clustering coefficient (significantly different between densities of 0.06-0.32) for BiTLE (green) and UTLE (red). Dashed lines represent the mean, shaded curve represents 95% CI. a.u. stands for arbitrary units. * p<0.05.

In a *post-*hoc analysis, we aimed to determine which nodes were driving the differences in average brain clustering between the two groups. **Figure 4C** shows the Brainnetome atlas regions that were found to have significantly different clustering between BiTLE and UTLE at densities of 0.2, 0.3 and 0.4. We chose these densities because they spanned the range where moderate effect sizes were observed for the clustering coefficient (**Supplementary Figure 3D**). Across the three studied densities, we found a high fraction of the regions in the default mode network (DMN), somatomotor network, salience ventral attention and frontoparietal network that differed significantly between groups (**Figure 4B,C**). These findings suggest that changes in clustering within multiple functional networks might contribute to the differences observed in the whole brain clustering coefficient between the two groups.

**Figure 4.**
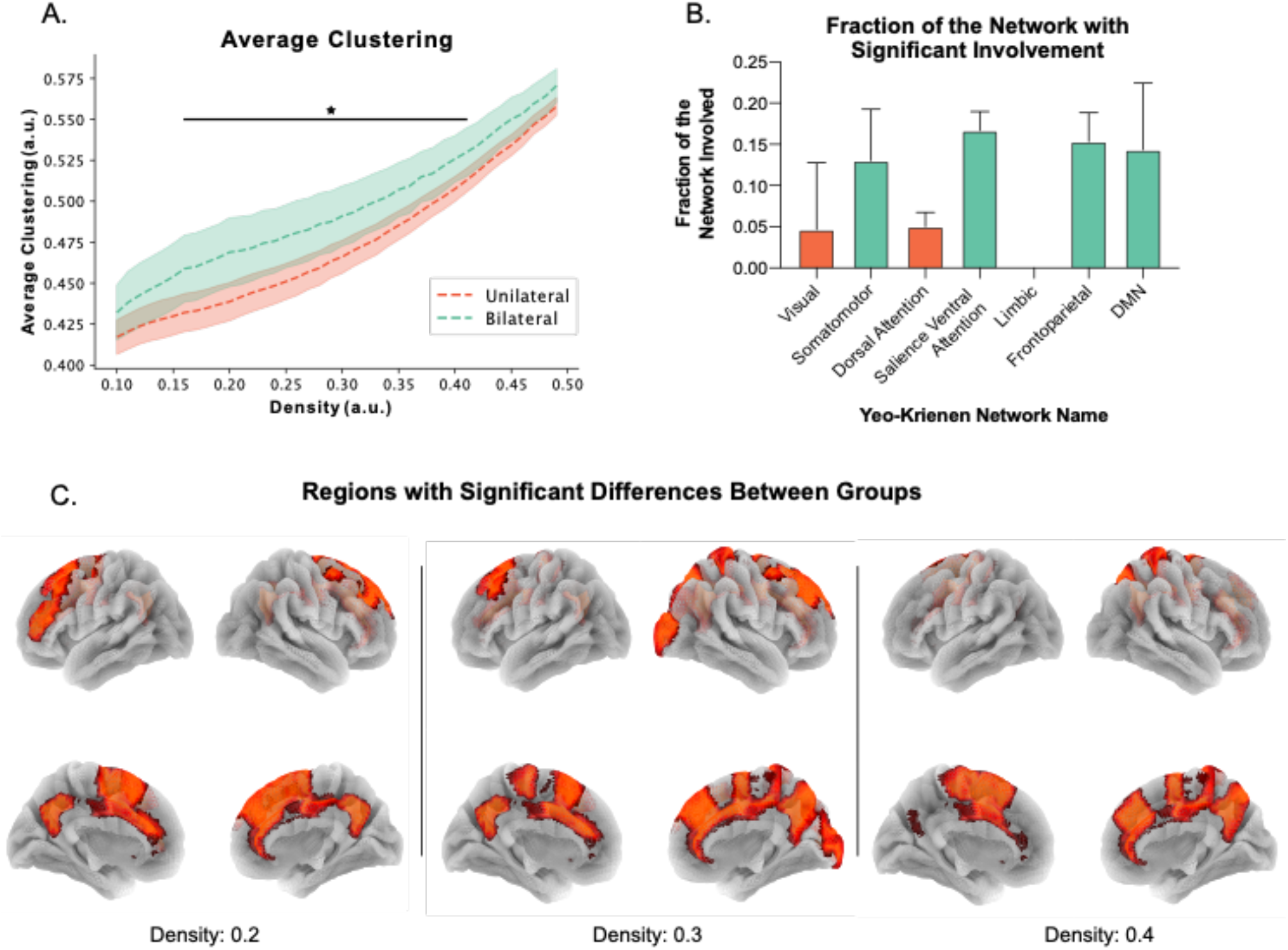
Nodal Clustering Coefficient: A. Whole-brain average clustering coefficient (repeat of Figure 1D). B. Fraction of the network with significantly different clustering coefficient (defined as number of regions with a significant difference divided by the total number of regions in the network) between BiTLE and UTLE at network densities of 0.2, 0.3, and 0.4. Bars represent the mean, and error bars represent 95% CI across the three densities. C. Regions with significantly different (p<0.05) clustering coefficient between UTLE and BiTLE at densities of 0.2, 0.3 and 0.4.

### Decreased Whole Brain and Temporal Participation in BiTLE

To assess the level of interaction among different brain communities in each patient group, we quantified the participation coefficient using communities defined by the boundaries of the lobes of the brain (anatomic parcellations), and parcellations defined by the functional networks of the brain (functional parcellations). The participation coefficient in functional and anatomic parcellations is shown in **Figure 5**. The average participation coefficient across the whole brain for the anatomic parcellations was not significantly different between the groups (p_cluster_=0.14). The average participation coefficient across the whole brain for the functional parcellations was significantly lower for the BiTLE group across the entire range of densities (p_cluster_=0.001).

**Figure 5.**
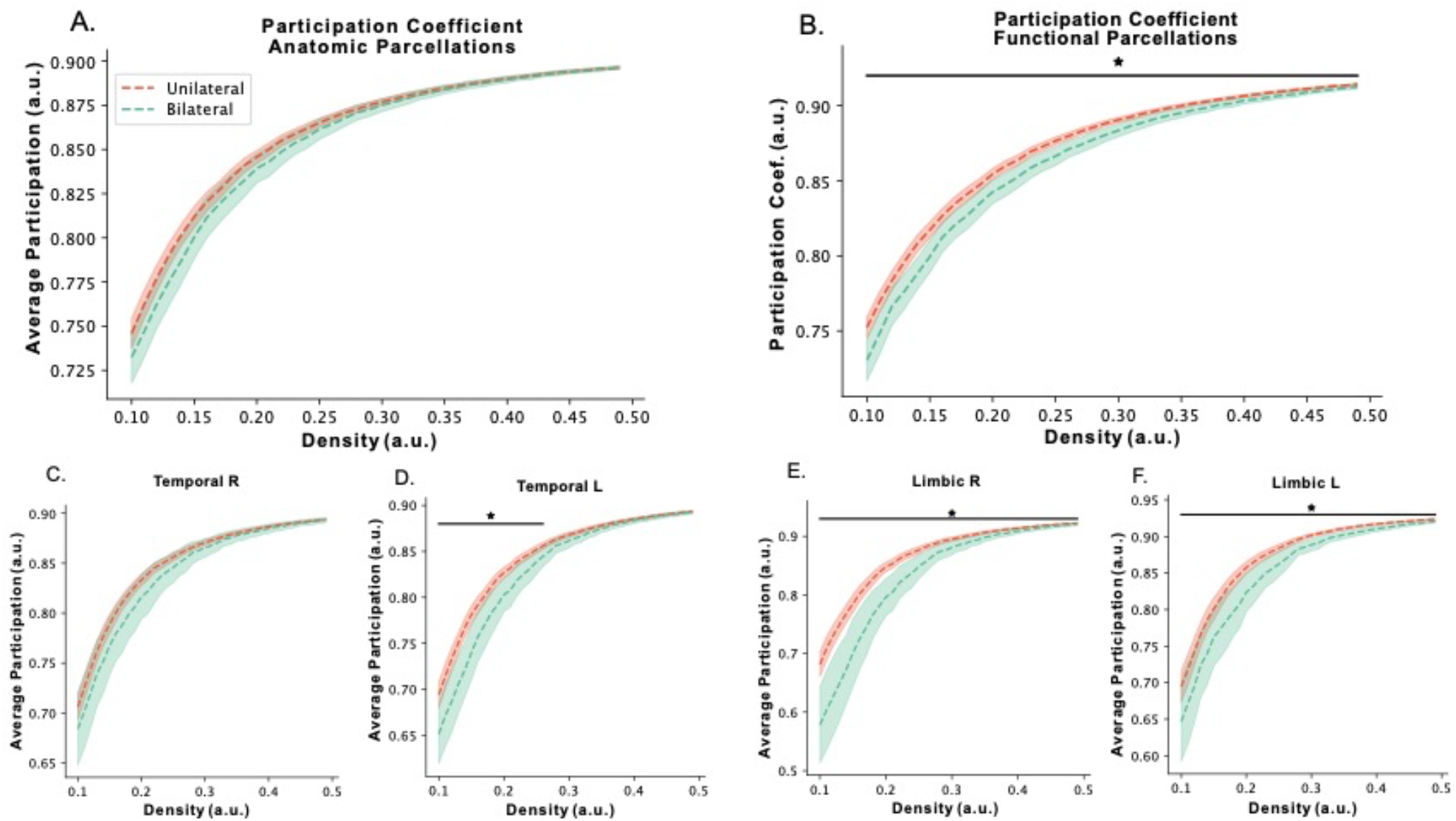
Participation Coefficient Results: Whole-brain average participation coefficient across densities for A. anatomically and B. functionally defined communities (significantly different across entire density range). Average participation coefficient for all nodes within the C. left/ipsilateral (significantly different between 0.10-0.27) and D. right/contralateral temporal community. Average participation coefficient for all nodes within the C. left/ipsilateral and D. right/contralateral Yeo-Krienen 7 limbic community assignment (both significantly different across entire density range). BiTLE (green) and UTLE (red). The Yeo-Krienen 7 limbic network consists of the basal frontal cortex and temporal poles. Dashed lines represent the mean, shaded curve represents 95% CI. a.u. stands for arbitrary units. * p<0.05

Inspecting the average participation coefficient within each of the anatomic lobes (**Figure 5C,D**) showed that there was significantly decreased participation within the temporal lobe for the ipsilateral hemisphere of UTLE and left hemisphere of BiTLE across densities of 0.10-0.27 (p_cluster_=0.026). No significant differences in participation coefficient were found between the contralateral UTLE and right BiTLE temporal lobe (p_cluster_=0.081), although the results trended in the same direction as in the opposite hemisphere. No significant differences were found in the other lobes (**Supplementary Figure 4**). Within functional communities, there was significantly lower average participation coefficient (p_cluster_=0.001) bilaterally in the “limbic” Yeo-Krienen system across the entire range of densities (**Figure 5E,F**). There was also significantly lower participation (p_cluster_=0.007) in the right BiTLE visual network compared to the contralateral UTLE visual network across all density ranges. No significant differences were found in the other functional communities (**Supplementary Figure 5**). Overall, our results suggest decreased average brain inter-community participation in BiTLE relative to UTLE, which might be driven by decreased inter-community participation of the temporal lobe.

### Increased Number of Smaller Communities Detected in BiTLE

As another way to test segregation and integration, we employed a modularity maximization procedure to determine, in a data driven way, the number of communities present in BiTLE and UTLE. In this analysis, a larger number of communities detected would correspond to a more segregated whole brain network. The results for the number of communities detected after modularity maximization are shown in **Figure 6**. For the density-dependent networks (γ=1), a significantly larger (p_cluster_=0.016) number of communities were detected for the BiTLE group in the density range between 0.10-0.14, but the opposite effect was observed between density ranges 0.42-0.50 (p_cluster_=0.016). That is, a larger number of communities were detected at smaller network sizes in BiTLE compared to UTLE. However, at larger network sizes, specifically between densities of 0.42-0.50, BiTLE was found to have a smaller number of communities. The quality of the modularity was not significantly different between the two groups, although the value was on average higher for the BiTLE group (**Supplementary Figure 6A**). Using the unthresholded functional connectome at different values of the resolution parameter γ, we found a significantly larger (p_cluster_=0.030) number of communities in BiTLE relative to UTLE at values of γ>2. That is, a larger number of smaller communities were detected in BiTLE than in UTLE at larger γ values. However, the quality of the modularity was exponentially lower for those larger values of γ in both groups (**Supplementary Figure 6B**). Overall, a larger number of smaller communities, both at low network density and at high γ, were detected in BiTLE.

**Figure 6.**
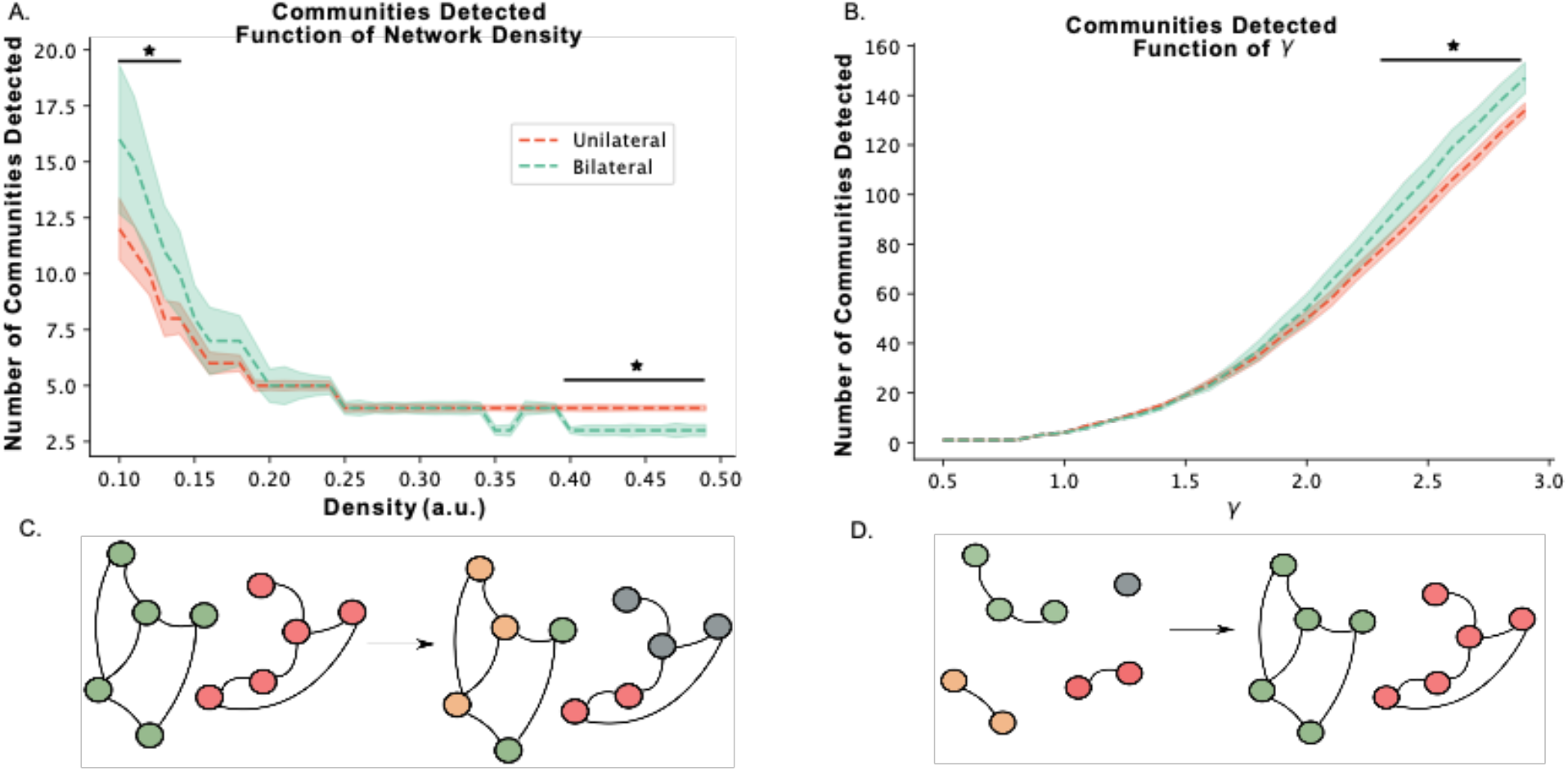
Modularity Maximization Results: Number of communities detected after modularity maximization in A. density dependent fashion while keeping ϒ=1 (significantly different at density values of 0.10-0.14, 0.42-0.50), B. unthresholded functional connectome as a function of increasing values of γ (significantly different at density values of 2.5-3.0), BiTLE (green) and UTLE (red). Dashed lines represent the mean, shaded curve represents 95% CI. a.u. stands for arbitrary units. * p<0.05. C. and D. are visual representations of the effects changing ϒ or network sparsity has on the communities detected through modularity maximization. C. Shows that for a fixed network size (i.e., the unthresholded functional connectome), as we increase γ, the size of the detected communities decreases, and since the number of connected nodes stays the same, number of communities detected has to increase. B. Shows that for a fixed ϒ, as the sparsity of the network decreases (or the density increases), larger communities are detected and therefore the number of communities detected decreases.

### The Integration-Segregation Axis Separates BiTLE from UTLE

We generated and integration-segregation axis through principal component analysis of the global efficiency and average clustering density curves of each subject. Spectral clustering of each subject in the principal component space resulted in two approximately equally sized clusters as shown in **Figure 7A**. One cluster was distinctly located on the higher segregation portion of the first principal component and the other cluster was located on the higher integration portion. **Figure 7B** shows the distribution of UTLE and BiTLE patients in the principal component space. From the BiTLE group, 68% (13/19) patients were identified within the high segregation cluster, whereas only 41% (31/75) of the UTLE patients were identified in the high segregation cluster (p=0.042, χ_^2^_=4.24). The distribution in the principal component space of UTLE patients that underwent surgical ablation/resection and had good (Engel score = I) or poor (Engel score > I) seizure surgical outcomes is shown in **Figure 7C**. From the poor surgical outcome group, 78% (7/9) of the patients were identified in the high segregation cluster, whereas only 35% (6/17) of the good surgical outcome patients were found in the high segregation cluster (p=0.09, Fischer’s exact test). We estimated whether the value of PC1 was higher for BiTLE and for poor outcome UTLE patients (**Figure 7D**). We found that PC1 was significantly higher for BiTLE relative to UTLE (p=0.026, two-sided *t-*test). We also found that PC1 was significantly higher for poor outcome UTLE relative to good outcome UTLE (p=0.028, two-sided *t*-test). We found no differences in the value of PC1 for poor outcome UTLE patients and BiTLE (p=0.72, two-sided *t*-test). Overall, both BiTLE and UTLE patients with poor post-surgical outcome were preferentially located in the high segregation portion of the integration-segregation axis.

**Figure 7.**
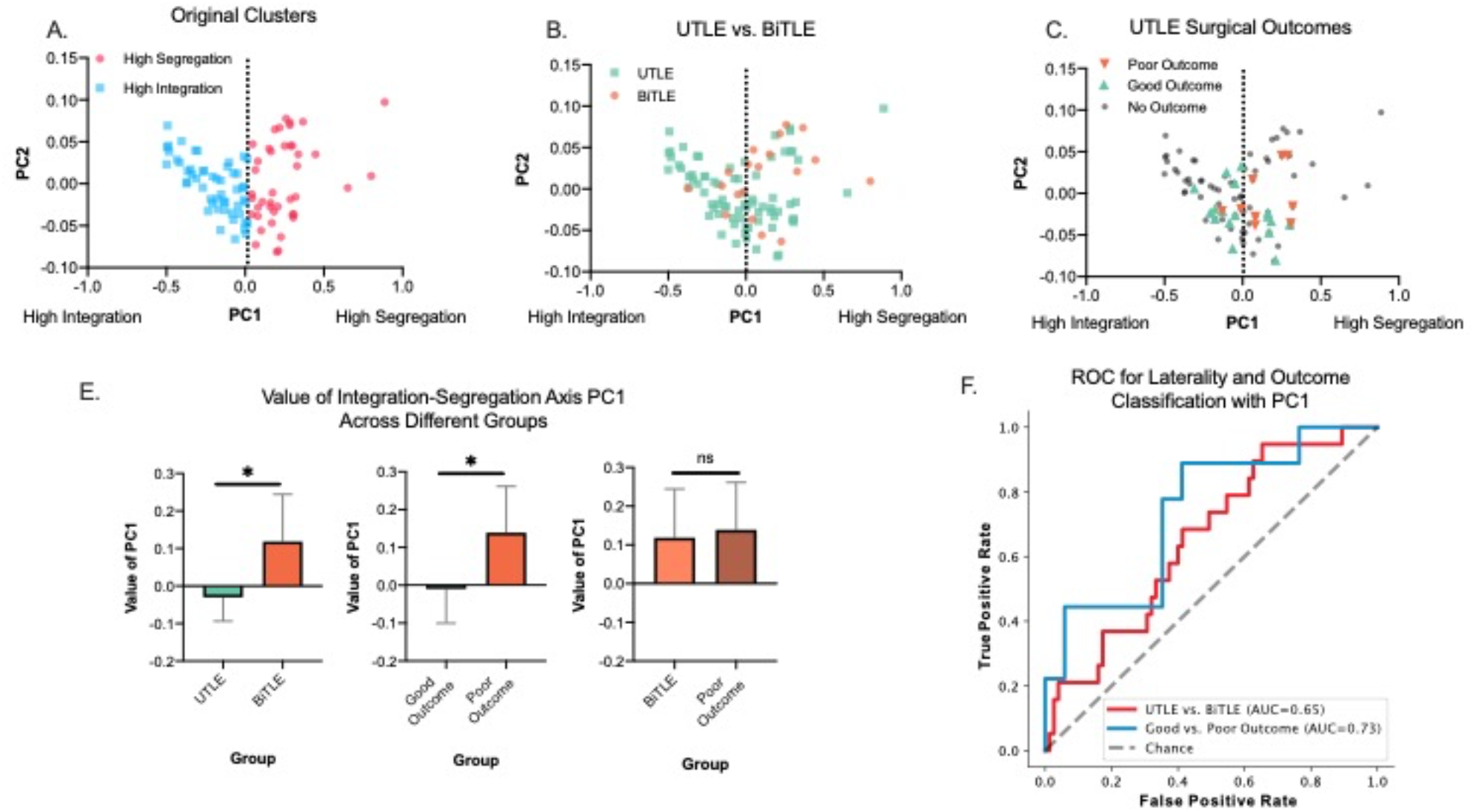
Integration-segregation axis results: **A**. Cluster assignments after spectral clustering. Dashed line represents the margin between the two detected clusters. **B**. Location of BiTLE and UTLE subjects along the first and second principal components of the integration segregation axis. **C**. Location of UTLE subjects with good and poor surgical outcomes along the first and second principal components of the integration segregation axis. Those without surgical outcomes reported are shown in gray. D. Value of PC1 (horizontal axis in figures A-C), for UTLE and BiTLE subjects, good and poor surgical outcome UTLE subjects, and BiTLE and poor outcome UTLE subjects. Bars represent the mean and error bars 95% CI. * p<0.05. E. Receiver operating characteristics curves for separating UTLE and BiTLE (red line), and good vs. poor outcome UTLE (blue line) using PC1 from the integration-segregation axis as a metric. Dashed line represents value for a random classifier. AUC: area under the curve.

Finally, to validate whether the value of PC1 would be capable of distinguishing UTLE from BiTLE, and good from poor outcome UTLE subjects, at a single subject level, we assessed its receiver operating characteristics (ROC) (**Figure 7E**). Overall, the area under the curve (AUC) was 0.65 for distinguishing UTLE from BiTLE, and 0.73 for distinguishing good and poor outcome UTLE subjects.

## Discussion

In this study, we aimed to characterize differences in resting state functional network organization in BiTLE and UTLE. Our results suggest an increase in brain segregation in BiTLE when compared to UTLE. The increased segregation was directly quantified by a decreased whole brain global efficiency and participation coefficient, as well as by an increased number of communities detected across the brain. Furthermore, we generated an integration-segregation axis from a linear combination of the average clustering coefficient and global efficiency across densities and found that BiTLE patients were mostly clustered in the high segregation portion of the spectrum, as were patients with UTLE that showed poor post-operative seizure outcomes. Increased segregation of the epileptic resting-state network supports prior work, as large meta-analytical studies have demonstrated increased interictal network segregation in patients with epilepsy when compared to neurotypical controls^31^. Our results provide novel insights by further indicating increased segregation in BiTLE relative to UTLE, and shed light on the underlying organization of the interictal network in BiTLE^6^.

Our findings suggest that changes in the DMN contribute to the increased segregation in BiTLE, particularly regarding the increased clustering coefficient of the network. This finding builds on prior evidence, as functional alterations of the DMN have been often implicated in epilepsy^59–61^. Particularly in UTLE, decreased ipsilateral hippocampal connectivity to the DMN has been attributed to decreased recruitment of the ipsilateral hippocampus by the DMN^62^. This could be interpreted as increased ipsilateral disconnection of either the hippocampus or the DMN. Given prior knowledge about DMN segregation in UTLE, it is reasonable to expect increased bilateral DMN segregation in BiTLE. In addition to the DMN, however, we also saw similar involvement of the salience ventral attention, frontoparietal, and somatomotor networks, all of which have also been associated with epilepsy^63,64^. In fact, a study of changes in structural connectivity in focal epilepsy^64^ found that in the studied sample, the most common functional networks involved were: salience, DMN, frontoparietal and somatomotor, in that order. Therefore, it is likely that an interaction across all these functional networks is what is driving the increased clustering observed in BiTLE.

The participation coefficient of brain regions has been used in the study of network segregation during development^65^, as well as in assessing the role of thalamocortical networks in bilateral tonic-clonic seizures^66^. Our results suggest that in BiTLE, the temporal lobe and frontotemporal area (Yeo-Krienen “limbic” system) are characterized by increased segregation, as quantified by their decreased intra-network participation coefficient. The bilaterally decreased temporal participation coefficient found in our study is consistent with the involvement of the bilateral temporal lobes in independently generating seizures in BiTLE. This may suggest that decreased temporal and frontotemporal integration in BiTLE contributes to a highly segregated network in the bilateral temporal lobes that drives epileptogenicity.

Decreased integration and increased segregation of the resting-state whole-brain functional network appears to be a pattern that correlates with disease complexity, at least in epilepsy. A recent study in pediatric febrile seizures demonstrated a very similar pattern of segregation-integration as the one in our study, when comparing complex and simple febrile seizures^47^: children with complex febrile seizures had increased average clustering, decreased global efficiency, and decreased average participation coefficient relative to those with simple febrile seizures. Furthermore, converging evidence supports the findings that people with epilepsy have higher clustering and lower global efficiency than healthy controls^19,31,67^. This increase in clustering has been observed in UTLE even when concurrent neuronal fiber loss has been reported^68^. This “paradoxical” strengthening of local connectivity despite fiber loss is possibly part of the mechanisms in TLE, where abnormal plasticity hijacks connectivity to local epileptogenic structures. Therefore, it is possible that the resting state functional network becomes progressively more segregated on the disease spectrum from a neurotypical network to focal epilepsy (i.e. UTLE), and from focal epilepsy to a potentially multifocal disease (BiTLE)^69^.

Increased network segregation in BiTLE, as demonstrated here, provides evidence for BiTLE being a multi focal disease, where areas of epileptogenesis may act as smaller independent segregated networks. Our findings are thus consistent with the current clinical definition of BiTLE, where both hemispheres are required to be seizing independently, and in contrast with the hypothesis that a primary focus may drive both hemispheres, which would be instead supported by a finding of more integrated network topology. The ‘independent foci’ hypothesis, in turn, could also explain why poor outcome UTLE subjects appear similar to BiTLE along the integration segregation axis. In the presence of more segregated networks, if a suspected epileptic focus is removed, there may still be a possibility of a secondary independent foci being active, causing the patient to continue seizing, even after surgical removal of the primary focus. At an individual subject level, however, we observe that there is indeed a distribution of subjects across the integration segregation axis, and there is likely a spectrum of multifocality that, to some extent, may apply to both UTLE and BiTLE.

Another hypothesis is that the increased interictal segregation in epilepsy is a counter-regulatory mechanism for suppressing seizure spread from the SOZ^19^. fMRI recordings in epilepsy occur interictally and patients are not assumed to be seizing during image acquisition. The inhibitory activity suppressing the ictal network could potentially be driving the resting state network into a more segregated state, preventing seizure spread by containing the epileptogenic regions. This hypothesis, in turn, would be consistent with increased segregation as disease severity and complexity increases, since a stronger counter-regulatory network would be required to suppress seizures in these instances. One way to further assess the validity of this argument is to obtain concurrent EEG during fMRI studies, as is routinely done at many centers for PET studies, in order confirm a truly interictal state.

The network differences between BiTLE and UTLE detected in this study could have significant clinical impact, particularly in the pre-surgical planning of epilepsy surgery.

Differentiating patients with BiTLE vs. UTLE has immense clinical value, as invasive procedures, such as intracranial electrode placement, are often required to make this distinction. In addition, even in patients who undergo invasive intracranial EEG monitoring, the relatively short duration of monitoring may not capture bilateral onset seizures^4^. Furthermore, with the increased use of RNS and DBS^70^ in the treatment of BiTLE, being able to identify candidate patients may allow clinicians to proceed to neuromodulatory therapy with greater confidence, displacing the burden of diagnosis solely from the results of iEEG. Given the non-invasiveness and widespread availability of fMRI, measurement of resting state topological properties could feasibly be incorporated into the presurgical planning pipeline. The integration segregation axis developed in this study provides a proof of concept for non-invasively identifying BiTLE and poor outcome UTLE patients during the presurgical evaluation using the results of this study. We obtained AUCs that were well above baseline for distinguishing BiTLE from UTLE as well as good and poor outcome UTLE patients, but we believe that more intricate models that make use of machine learning in combination with our results might provide further insight as well as better classification performance. We provide all the processed data used in this study, as well as the source code for each part of the analysis, as an incentive for other researchers to continue exploring the differences between BiTLE and UTLE. In future work, we envision a platform enabling clinicians access to our metric estimating the likelihood of BiTLE vs. UTLE and overall surgical outcome. Ideally, such a platform would utilize artificial intelligence strategies (e.g., machine learning) with multiple quantitative clinical and imaging features to generate these predictions at the individual patient level.

Our study has limitations. First, despite the relatively large sample size, the BiTLE group was somewhat underrepresented relative to the UTLE group. This is a consequence of both the relative decreased incidence of BiTLE but also the complexity of this condition, which can lead to potential misclassification of a truly BiTLE patient as UTLE. However, as academic centers expand their surgical epilepsy programs and our presurgical pipelines become more robust, more BiTLE patients are likely to be identified. Second, our study does include patients from two different institutions, but the BiTLE cohort in this study was from only one of the two institutions. One of the strengths of our approach is the use of NeuroCombat to harmonize data across centers, minimizing the risk of site-dependent effects. Future studies should aim at increasing the sample size of BiTLE patients, while also including subjects from multiple institutions. Third, our study primarily focused on characterizing interictal resting state networks with fMRI. While appropriate due to fMRI’s whole brain coverage and non-invasiveness, the temporal resolution is poor compared to more invasive intracranial EEG approaches. Future studies will seek to better understand the interictal network of BiTLE using both neuroimaging as well as intracranial recordings. Finally, our study did not show significant findings in the subcortical structures (i.e. hippocampus, thalamus, basal ganglia, etc.), which are known to be heavily involved in epilepsy^20,66,71^. A potential explanation for this is that the low signal-to-noise ratio of fMRI in subcortical regions leads to lower effect sizes within subcortical structures, which are usually not sufficient for surviving multiple comparison correction^72–75^. Alternatively, it could be that UTLE and BiTLE only differ cortically and not subcortically. Future studies should aim to study the hippocampal, thalamic, and basal ganglia networks through a more directed and hypothesis-driven strategy in order to better characterize these networks in BiTLE.

### Conclusion

We found increased interictal resting state network segregation in BiTLE compared to UTLE as quantified by decreased global efficiency, increased clustering, decreased participation and increased number of communities detected. Differences in network properties place BiTLE and UTLE at different points in the integration-segregation spectrum, with higher segregation in BiTLE. UTLE patients with poor surgical outcomes are more similar to BiTLE than those with good surgical outcomes, also having potentially higher segregation. Our findings suggest that increased interictal whole brain network segregation, as measured by rs-fMRI, is specific to BiTLE, and may assist in non-invasively identifying this patient population prior to intracranial EEG or device implantation.

## Data Availability

https://doi.org/10.26275/t8ls-zuw2

