## Supplementary Methods and Figures for "Resting State Functional Connectivity Demonstrates Increased Segregation in Bilateral Temporal Lobe Epilepsy"

#### Subject Demographics

Data acquisition for this study was approved by the institutional review boards of the University of Pennsylvania (Penn) and the University of Liverpool (UOL). We compiled a multi-center dataset consisting of 94 drug-resistant TLE patients who had undergone neuroimaging during presurgical evaluation including resting state functional magnetic resonance imaging (fMRI).

From Penn, we studied 77 patients with drug resistant temporal lobe epilepsy who were in various stages of evaluation, including medical management, Phase I monitoring, phase II monitoring, and post-interventional (including resection, laser ablation, and stimulation devices). All patients had advanced imaging and final localization in the temporal lobe, which was indicated by the final localization determined by the Penn Epilepsy Surgical Conference (PESC Lateralization and PESC Localization) after evaluation of various clinical factors, which could include seizure semiology, MRI, PET, MEG, scalp EEG, and intracranial EEG. Thirty-four patients had left-sided lateralization, 24 patients had right-sided lateralization, and 19 patients were bilateral. From a demographic perspective, the patient population was noted to be 54.5% female with a mean age of 38 $\pm$ 12.9. Thirty-one patients underwent surgical intervention, with 7 Engel IA, 9 Engel IB, 1 Engel IIA, 6 Engel IIB, 1 Engel IID, 5 Engel IIIA, 1 Engel IVB, and 1 Engel IVC; of the 9 implanted patients, 3 had VNS, 4 had RNS, and 2 had DBS. For the patients with BiTLE, 6 underwent surgical intervention, with 3 of them having RNS placement and the other 3 undergoing surgical resection. The 3 BiTLE patients that underwent surgical resection were deemed to have bilateral seizure onset upon further investigation after lack of post-surgical improvement in seizure outcome.

The MRI and EEG results are documented in the **Supplementary Table 1** to help delineate the localization decision. MRI lesion status (“MRI”), lesion lateralization (“MRI Lateralization”), and lesion localization (“MRI Localization”) were determined based on the final radiologic impression of the Penn Epilepsy Surgical Conference (PESC). There were 46 patients with lesions seen on MRI, of which 24 had left-sided lateralization, 16 had right-sided lateralization, and 6 were bilateral. Forty-two patients had lesions in the temporal lobe, while 4 patients were found to have lesions that either included another region (3), or solely the frontal lobe (1).

EEG Lateralization and EEG Localization were also determined based on the final impression of the Penn Epilepsy Surgical Conference (PESC). There were 34 patients who had at most intracranial EEG, for which intracranial EEG lateralized 17 to the left, 8 to the right, and 9 bilaterally. Of these 34 intracranial EEG patients, 30 were localized solely to the temporal lobe, while 3 included both frontal and temporal lobes and 1 included the frontal, temporal, and parietal lobes. There were 43 patients who only had scalp EEG (consisting of extended ambulatory monitoring or phase I monitoring), for which 17 patients were left-lateralized, 16 were right-lateralized, and 10 were bilateral. Of these 43 scalp EEG patients, 37 were localized solely to the temporal lobe, while 6 were localized to both frontal and temporal lobes.

From the UOL, an additional cohort of 17 drug resistant TLE patients was included. Localization and seizure focus was determined through a multidisciplinary surgical conference as with the Penn Cohort. All the patients in this cohort were unilateral TLE patients with 11 left-sided, and 6 right-sided seizure onset (SOZ) lateralization. Age, gender, MRI lesional status, and final lateralization are summarized in **Table 1.**

All subject identifiers included in **Supplementary Table 1** were randomly generated and were only known to the research group.

### Neuroimaging Processing

Preprocessing was performed using fMRIPrep 20.2.3 [1], which is based on Nipype 1.6.1[2].

#### *Anatomical data preprocessing*

For each subject, T1w images were corrected for intensity non-uniformity (INU) with N4BiasFieldCorrection[3], distributed with ANTs 2.3.3[4]. The T1w-reference was then skull-stripped with a Nipype implementation of the antsBrainExtraction.sh workflow (from ANTs), using OASIS30ANTs as target template. Brain tissue segmentation of cerebrospinal fluid (CSF), white-matter (WM) and gray-matter (GM) was performed on the brain-extracted T1w using FAST (FSL 5.0.9)[5]. A T1w-reference map was computed after registration of 2 T1w images (after INU-correction) using mri_robust_template (FreeSurfer 6.0.1)[6]. Brain surfaces were reconstructed using FreeSurfer's recon-all, and the brain mask estimated previously was refined with a custom variation of the method to reconcile ANTs-derived and FreeSurfer-derived segmentations of the cortical gray-matter of Mindboggle[7]. Volume-based spatial normalization to one standard space (MNI152NLin2009cAsym) was performed through nonlinear registration with antsRegistration (ANTs 2.3.3), using brain-extracted versions of both T1w reference and the T1w template. The following template was selected for spatial normalization: ICBM 152 Nonlinear Asymmetrical template version 2009c [8].

#### *Functional data preprocessing*

For the single BOLD run of each subject, the following preprocessing was performed. First, a reference volume and its skull-stripped version were generated using a custom methodology of fMRIPrep. A B0-nonuniformity map (or fieldmap) was estimated based on a phase-difference map calculated with a dual-echo GRE (gradient-recall echo) sequence, processed with a custom workflow of SDCFlows inspired by the epidewarp.fsl script and further improvements in HCP Pipelines[9]. The fieldmap was then co-registered to the target EPI (echo-planar imaging) reference run and converted to a displacements field map (amenable to registration tools such as ANTs) with FSL's fugue and other SDCflows tools. Based on the estimated susceptibility distortion, a corrected EPI (echo-planar imaging) reference was calculated for a more accurate co-registration with the anatomical reference. The BOLD reference was then co-registered to the T1w reference using bbregister (FreeSurfer) which implements boundary-based registration[10]. Co-registration was configured with six degrees of freedom. Head-motion parameters with respect to the BOLD reference (transformation matrices, and six corresponding rotation and translation parameters) are estimated before any spatiotemporal filtering using mcflirt (FSL 5.0.9)[11]. BOLD runs were slice-time corrected using 3dTshift from AFNI 20160207[12]. The BOLD time-series (including slice-timing correction when applied) were resampled onto their original, native space by applying a single, composite transform to correct for head-motion and susceptibility distortions. These resampled BOLD time-series will be referred to as preprocessed BOLD in original space, or just preprocessed BOLD. The BOLD time-series were resampled into standard space, generating a preprocessed BOLD run in MNI152NLin2009cAsym space. First, a reference volume and its skull-stripped version were generated using a custom methodology of fMRIPrep. Several confounding time-series were calculated based on the preprocessed BOLD: framewise displacement (FD), DVARS and three region-wise global signals. FD was computed using two formulations following Power (absolute sum of relative motions)[13] and Jenkinson (relative root mean square displacement between affines)[11]. FD and DVARS are calculated for each functional run, both using their implementations in Nipype[13]. The three global signals are extracted within the CSF, the WM, and the whole-brain masks. Additionally, a set of physiological regressors were extracted to allow for component-based noise correction (CompCor)[14]. Principal components are estimated after high-pass filtering the preprocessed BOLD time-series (using a discrete cosine filter with 128s cut-off) for the two CompCor variants: temporal (tCompCor) and anatomical (aCompCor). tCompCor components are then calculated from the top 2% variable voxels within the brain mask. For aCompCor, three probabilistic masks (CSF, WM and combined CSF+WM) are generated in anatomical space. The implementation differs from that of Behzadi et al.[14] in that instead of eroding the masks by 2 pixels on BOLD space, the aCompCor masks are subtracted a mask of pixels that likely contain a volume fraction of GM. This mask is obtained by dilating a GM mask extracted from the FreeSurfer's aseg segmentation, and it ensures components are not extracted from voxels containing a minimal fraction of GM. Finally, these masks are resampled into BOLD space and binarized by thresholding at 0.99 (as in the original implementation). Components are also calculated separately within the WM and CSF masks. For each CompCor decomposition, the k components with the largest singular values are retained, such that the retained components' time series are sufficient to explain 50 percent of variance across the nuisance mask (CSF, WM, combined, or temporal). The remaining components are dropped from consideration. The head-motion estimates calculated in the correction step were also placed within the corresponding confounds file. The confound time series derived from head motion estimates and global signals were expanded with the inclusion of temporal derivatives and quadratic terms for each (Satterthwaite et al. 2013). Frames that exceeded a threshold of 0.5 mm FD or 1.5 standardised DVARS were annotated as motion outliers. All resamplings can be performed with a single interpolation step by composing all the pertinent transformations (i.e. head-motion transform matrices, susceptibility distortion correction when available, and co-registrations to anatomical and output spaces). Gridded (volumetric) resamplings were performed using antsApplyTransforms (ANTs), configured with Lanczos interpolation to minimize the smoothing effects of other kernels (Lanczos 1964). Non-gridded (surface) resamplings were performed using mri_vol2surf (FreeSurfer).

The output of the fMRIPrep pre-processing pipeline was subsequently input into the xcpEngine post-processing pipeline[15]. The xcpEngine post-processing pipeline is a self-contained software that allows the rapid and reproducible implementation of tools necessary for calculating functional connectivity, while also allowing for benchmarking pipeline performance using a wide array of benchmarking pipelines[16]. xcpEngine is built to use the output of the fMRIPrep pipeline as an input, therefore, many of the pipeline steps use metrics explicitly calculated by fMRIPrep. Briefly, the steps implemented in the xcpEngine pipeline for each subject, were as follows. First, regressors for artifactual signals were calculated from the 4D time series of each subject using the *confound2* module. The regressed parameters calculated by this module included motion realignment parameters (3 rotational and 3 translational) necessary for realigning each volume in the time series to a reference volume; the mean white matter and cerebrospinal fluid time series over all voxels[17], with tissue segmentations determined by the fMRIPrep; the mean time series signal across the whole brain[13]; the temporal derivative of motion parameters, which encodes the relative displacement of the brain from one volume of the timeseries to the next[17]; and finally, the second power of each of the previously mentioned regressors was also included, to account for potential noise that is proportional to higher powers of motion and nuisance regressors (a total of 36 regression parameters). After estimating the regressors, demeaning and detrending, followed by temporal filtering was carried out in both the BOLD timeseries and the regressors, using the *regress* module. The timeseries and regressors were detrended using a 2$^{nd}$ order polynomial. A first order forward-backward bandpass Butterworth filter, with passband 0.01-0.10Hz was implemented, allowing both high frequency noise, and very-low-frequency drift to be eliminated. The filtered regressors were fitted to the filtered BOLD timeseries data using multiple linear regression. Any variance in the BOLD timeseries explained by the regressors was discarded from the timeseries, whereas the unexplained variance was left as the final filtered timeseries.

**Network Metrics**

We assessed the topological properties of the functional connectivity matrices in a connection-density-dependent manner, to avoid the use of an arbitrary threshold[18,19–22]. Network topological properties were estimated at proportional thresholds representing connection densities ranging from 10% to 50%, with 1% increments[23]. During thresholding, the absolute value of all connections was taken. We chose the lower bound of 10% since networks fragment at low densities, and the upper bound of 50% since network properties at higher densities tend to randomness due to the inclusion of potentially confounding associations[19,20].

Across the network density range specified above, we computed the following metrics: degree, betweenness centrality, clustering coefficient and global efficiency. The degree quantifies the number of connections present in any given node. Betweenness centrality quantifies the amount of influence a node has over the flow of information in the network by measuring how much of a "bridge" a node is by separating two parts of the network. Finally, the clustering coefficient and global efficiency provide two direct measurements of network integration and segregation. The clustering coefficient quantifies the extent to which neighbors of a given node are neighbors of each other, serving as a direct measurement of segregation[24]. On the other hand, global efficiency quantifies how easily any given node can reach any other node in the network, serving as a direct measurement of integration[25].

The degree, betweenness centrality, eigenvector centrality, and clustering coefficient were computed for each node (or parcellation), and subsequently averaged across all nodes in the network, resulting in a single value for each metric across densities. The resulting density curves allow for the comparison of each averaged network metric at different levels of network sparsity.

### Participation Coefficient

The participation coefficient determines the degree to which a node that belongs to a given community interacts with nodes of other communities. Communities that have a high participation coefficient have connections to many other communities, whereas those communities with a low participation coefficient have few connections to other communities. In other words, communities with high participation coefficients are less segregated than communities with lower participation coefficients [26,27]. The participation coefficient of brain regions has been used in the study of network segregation during development [28], as well as in assessing the role of thalamocortical networks in bilateral tonic-clonic seizures[29].

In this study the participation coefficient, initially proposed in [27], was defined as:

$$P_i = 1 - \sum_{s=1}^{N_M} \left(\frac{\kappa_{i,s}}{k_i}\right)^2$$

Where $P_i$ is the participation coefficient of node $i$, $\kappa_{i,s}$ is the number of links from node $i$ to nodes in community $s$, and $k_i$ is the degree of node $i$. In this quantity, values closer to 1 indicate that a given node has connections uniformly distributed across all the communities, whereas values closer to 0 indicate that a given node has connections exclusively within a single community.

We computed the participation coefficient[27] to directly assess the degree of integration/segregation of putative functional and anatomic brain communities. Functional community assignments provide a natural framework for studying participation across communities, since these assignments are based on rs-fMRI data[30]. Anatomic community assignments are useful in this setting since localizing network abnormalities within a specific anatomic location (e.g. temporal lobe in TLE) is of particular interest in epilepsy.

For the functional community assignments, the Yeo-Krienen 7 cortical networks were used[30]. Each of the 246 parcellations in the Brainnetome atlas was assigned to one of the 7 functional communities as provided by the atlas' documentation. The anatomic community assignments included: frontal, parietal, temporal, insular, occipital and limbic lobes, as well as a subcortical community, all of which were defined by the lobar assignments of the Brainnetome atlas. Both the anatomic and functional assignments were further split into left/right, for the BiTLE subjects, and ipsilateral/contralateral for the UTLE subjects. As with the other network metrics, the participation coefficient was computed at each node and then subsequently averaged across the entire brain. This process was repeated across the same range of densities defined previously.

Finally, to assess the contribution of each community to the whole brain average participation, the average participation of all nodes within each community, instead of across the whole brain, was also calculated.

**Modularity Maximization**

Modularity maximization was also carried out to identify the number of communities that could independently be detected for each subject, with the assumption that a network where a larger number of communities is detected is one with a higher degree of segregation. The Louvain algorithm for modularity maximization was utilized[31]. Under this approach, a community is defined as a set of highly interconnected nodes, which are identified through maximization of the modularity function, Q. Q measures the quality of the communities by measuring the density of connections inside as compared to the density of connections between communities[32]. Q is defined as:

$$Q = \frac{1}{2m}\sum_{i,j}\left(A_{i,j} - \gamma\frac{k_i k_j}{2m}\right)\delta(i,j)$$

Where $A$ is the adjacency matrix between nodes. $k_i$ and $k_j$ correspond to the sum of the weights assigned to nodes $i$ and $j$, respectively, and $m$ is the sum of all the edges within the graph. Overall, the quantity $\frac{k_i k_j}{2m}$ represents the expected weight of the connection between nodes $i$ and $j$, serving as a null connectivity model. Quantity $\delta(i,j)$ is the Kronecker delta, which is 1 if $i = j$. Finally, there is a resolution parameter, $\gamma$, which specifies the relative importance of the null model. Larger values of Q are taken to indicate higher quality partitions and represent communities with a larger number of intra-community connections than inter-community connections.

The resolution parameter $\gamma$ determines the size of the communities that are detected, where larger values of $\gamma$ lead to smaller communities. Given that the communities detected are different for each subject, the number of communities detected was the metric of interest. The community detection was performed in two ways. First, we performed community detection in a network density dependent manner, with the commonly used resolution parameter of $\gamma$=1. Second, we identified communities in the unthresholded functional connectome. This was done at a range of $\gamma$ values between 0.5-

3.0 with a spacing of 0.01 in order to detect the communities in a scale dependent way, as we expected that divergence between the two groups to happen in a scale-dependent manner[33]. The first approach quantifies how the number of communities detected in the two groups differ as the size of the network itself gets bigger (higher densities, larger network). The second approach quantifies how the number of communities detected in BiTLE and UTLE diverge as the detected communities get smaller (larger gamma, smaller communities).

**Statistical Analysis**

Statistical comparison of the network density dependent curves between BiTLE and UTLE was done through a 1-dimensional suprathreshold cluster detection procedure, analogous to the one proposed Nichols et. al. [34]. This approach is commonly used in the fMRI literature, and provides family wise error rate (FWER) correction without sacrificing too much power. Briefly, for a given network metric at each density, a Welch's *t*-test was carried out, generating a *t*-statistic at each density. This 1-dimensional vector of t-statistics was then thresholded at a significance level of $p<0.05$, uncorrected. Densities above the threshold were identified, and 1-dimensional clusters, consisting of groups of adjacent densities that surpassed the threshold, were defined. Then, the group labels for each subject were permuted 1000 times, and the process above was repeated for each permutation, generating 1000 different sets of 1-dimensional clusters. From the permuted clusters, a distribution consisting of the maximum cluster size in each permutation was generated. Cluster sizes from the unpermuted data that were larger than 95% of the maximum permuted cluster sizes were considered significant (equivalent to $p<0.05$, corrected). The p-value of this significant cluster is then estimated as the proportion of permuted cluster sizes that are larger than the largest cluster size in the unpermuted data, with the addition of a positive bias[35]. This procedure is well suited for our specific application for the following reasons. First, the differences in fMRI seen in epilepsy have small effect sizes[36], which requires the use of strategies that can increase statistical power, particularly in small sample sizes. Second, we are interested in answering the question of whether there is a range of network densities in which the measured network properties differ between the groups, and not necessarily at which specific densities this happens. The above clustering procedure increases statistical power, but it does so at the expense of only determining whether a cluster of densities is significant or not, and not which specific density is significant. For each metric, we also measured the absolute value of the Cohen's D between the two groups at each density to quantify effect sizes. We considered Cohen's D values above 0.4 to represent a moderate effect size, and above 0.6 to represent a large effect size.

In order to identify which nodes of the network had significant differences between the groups for a given density and network metric, we also applied a permutation-based cluster-wise procedure analogous to those used in past studies[19,37]. We applied this approach to nodes defined by the Brainnetome atlas, although we note that this procedure could be expanded to any atlas. Briefly, a binary spatial adjacency matrix A was generated, where rows and columns consisted of each region, and the values of the matrix were 1 if the two regions were spatially connected (i.e., atlas parcellations were right next to each other) and a value of 0 otherwise. Then, a Welch's *t*-test between the two groups was carried out at each parcellation, generating a t-statistic at each region. These *t*-statistics were thresholded at a value of $p<0.05$, and clusters of connected significant regions, as determined by A, were identified. As with the 1-dimensional case, 1000 permutations were obtained, and original clusters larger than 95% of the permuted clusters ($p<0.05$), were considered significant (**Supplementary Figure 2**). In these comparisons, regions in the left hemisphere of BiTLE were compared with regions in the ipsilateral hemisphere of UTLE, and regions in the right hemisphere of BiTLE were compared to regions in contralateral hemisphere of UTLE.

**Supplementary Table 1:** Detailed information for all subjects included in the study. Lesional: MRI lesional status. MRI Lat: Lateralization per MRI read. EEG: Type of EEG assessment. EEG Lat: Lateralization per EEG read. EEG Loc: Localization per EEG read. Final Lat: Final lateralization per multidisciplinary surgical conference. Final Loc: Final localization per multidisciplinary surgical conference. Surgical Outcome: Engel seizure surgical outcome score. Device: type of device implanted, if any. MTS: Presence of mesial temporal sclerosis.

| Subject | Cohort | Sex | Age Range | Lesional | MRI Lat | MRI Loc | EEG | EEG Lat | EEG Loc | Final Lat | Final Loc | Surgical Outcome | Device | MTS |
|---|---|---|---|---|---|---|---|---|---|---|---|---|---|---|
| sub-PENN01 | PENN | F | 16-20 | Lesional | R | Temporal | Intracranial | R | Temporal | R | Temporal | IIB | VNS | Y |
| sub-PENN02 | PENN | F | 16-20 | Lesional | R | Temporal | Intracranial | R | Frontal; Temporal | R | Temporal | IB | No | Y |
| sub-PENN03 | PENN | M | 51-55 | Lesional | B | Temporal | Intracranial | L | Temporal | L | Temporal | | No | |
| sub-PENN04 | PENN | M | 21-25 | Nonlesional | | | Intracranial | R | Temporal | R | Temporal | IB | No | |
| sub-PENN05 | PENN | F | 46-50 | Nonlesional | | | Intracranial | R | Temporal | R | Temporal | IB | No | |
| sub-PENN06 | PENN | F | 31-35 | Nonlesional | | | Intracranial | B | Frontal; Temporal | B | Temporal | | VNS | |
| sub-PENN07 | PENN | F | 26-30 | Nonlesional | | | Intracranial | L | Temporal | L | Temporal | IA | No | |
| sub-PENN08 | PENN | M | 31-35 | Nonlesional | | | Intracranial | R | Temporal | R | Temporal | IA | No | |
| sub-PENN09 | PENN | F | 46-50 | Lesional | L | Temporal | Intracranial | L | Temporal | L | Temporal | IB | No | |
| sub-PENN10 | PENN | F | 56-60 | Nonlesional | | | Intracranial | B | Temporal | B | Temporal | IIIA | RNS | |
| sub-PENN11 | PENN | F | 46-50 | Lesional | R | Temporal | Scalp | R | Temporal | R | Temporal | IA | No | |
| sub-PENN12 | PENN | M | 36-40 | Nonlesional | | | Intracranial | B | Temporal | B | Temporal | IVC | No | |
| sub-PENN13 | PENN | M | 41-45 | Nonlesional | | | Scalp | B | Temporal | B | Temporal | | No | |
| sub-PENN14 | PENN | F | 46-50 | Lesional | L | Temporal | Intracranial | L | Temporal | L | Temporal | IB | No | Y |
| sub-PENN15 | PENN | F | 41-45 | Nonlesional | | | Intracranial | B | Temporal | B | Temporal | | No | |
| sub-PENN16 | PENN | M | 46-50 | Lesional | R | Temporal | Scalp | B | Temporal | B | Temporal | IIIA | No | |
| sub-PENN17 | PENN | M | 21-25 | Lesional | L | Temporal | Intracranial | L | Temporal | L | Temporal | IB | No | Y |
| sub-PENN18 | PENN | F | 36-40 | Lesional | L | Temporal | Scalp | R | Temporal | R | Temporal | IIIA | VNS | Y |
| sub-PENN19 | PENN | M | 31-35 | Nonlesional | | | Intracranial | B | Temporal | B | Temporal | | No | |
| sub-PENN20 | PENN | F | 31-35 | Lesional | L | Temporal | Intracranial | L | Temporal | L | Temporal | IIIA | DBS | |

| | | | | | | | | | | | | | |
|---|---|---|---|---|---|---|---|---|---|---|---|---|---|
| sub-PENN21 | PENN | F | 41-45 | Lesional | L | Temporal | Scalp | B | Temporal | B | Temporal | | No | |
| sub-PENN22 | PENN | F | 46-50 | Lesional | L | Temporal | Scalp | L | Temporal | L | Temporal | IA | No | |
| sub-PENN23 | PENN | F | 21-25 | Nonlesional | | | Scalp | L | Temporal | L | Temporal | IA | No | |
| sub-PENN24 | PENN | F | 46-50 | Lesional | R | Temporal | Scalp | R | Temporal | R | Temporal | | No | |
| sub-PENN25 | PENN | M | 21-25 | Lesional | L | Temporal | Scalp | L | Frontal; Temporal | L | Temporal | | No | |
| sub-PENN26 | PENN | F | 31-35 | Lesional | L | Temporal | Intracranial | B | Temporal | B | Temporal | IIB | No | |
| sub-PENN27 | PENN | M | 31-35 | Nonlesional | | | Scalp | R | Temporal | R | Temporal | | No | |
| sub-PENN28 | PENN | M | 26-30 | Nonlesional | | | Intracranial | L | Frontal; Temporal; Parietal | L | Temporal | | No | |
| sub-PENN29 | PENN | F | 26-30 | Lesional | B | R cingulate, L temporal | Intracranial | L | Temporal | L | Temporal | IIB | No | |
| sub-PENN30 | PENN | F | 56-60 | Nonlesional | | | Scalp | R | Temporal | R | Temporal | | No | |
| sub-PENN31 | PENN | F | 21-25 | Lesional | B | Temporal | Intracranial | B | Temporal | B | Temporal | IIB | RNS | |
| sub-PENN32 | PENN | M | 21-25 | Lesional | L | Temporal; Insula | Intracranial | L | Temporal | L | Temporal | IA | No | |
| sub-PENN33 | PENN | F | 66-70 | Lesional | R | Temporal; Frontal | Scalp | R | Frontal; Temporal | R | Temporal | | No | Y |
| sub-PENN34 | PENN | M | 31-35 | Lesional | L | Temporal | Scalp | L | Temporal | L | Temporal | | No | |
| sub-PENN35 | PENN | M | 26-30 | Lesional | L | Temporal | Intracranial | L | Temporal | L | Temporal | | No | |
| sub-PENN36 | PENN | M | 21-25 | Nonlesional | | | Intracranial | R | Temporal | R | Temporal | | No | |
| sub-PENN37 | PENN | F | 26-30 | Nonlesional | | | Scalp | L | Temporal | L | Temporal | | No | |
| sub-PENN38 | PENN | F | 26-30 | Nonlesional | | | Intracranial | B | Temporal | B | Temporal | | No | |
| sub-PENN39 | PENN | F | 26-30 | Nonlesional | | | Scalp | B | Temporal | B | Temporal | | No | |
| sub-PENN40 | PENN | F | 31-35 | Lesional | L | Temporal | Scalp | L | Temporal | L | Temporal | | No | |
| sub-PENN41 | PENN | M | 31-35 | Lesional | L | Temporal | Intracranial | L | Temporal | L | Temporal | | No | |
| sub-PENN42 | PENN | M | 61-65 | Lesional | B | Temporal | Scalp | B | Temporal | B | Temporal | | No | |
| sub-PENN43 | PENN | F | 46-50 | Nonlesional | | | Intracranial | R | Frontal; Temporal | R | Temporal | IID | No | |
| sub-PENN44 | PENN | F | 46-50 | Lesional | L | Temporal | Scalp | L | Temporal | L | Temporal | | No | Y |
| sub-PENN45 | PENN | M | 51-55 | Nonlesional | | | Scalp | L | Temporal | L | Temporal | | No | |
| sub-PENN46 | PENN | F | 36-40 | Nonlesional | | | Intracranial | L | Temporal | L | Temporal | IB | RNS | |
| sub-PENN47 | PENN | M | 26-30 | Lesional | L | Temporal | Scalp | L | Temporal | L | Temporal | IIA | No | |

| | | | | | | | | | | | | | | |
|---|---|---|---|---|---|---|---|---|---|---|---|---|---|---|
| sub-PENN48 | PENN | M | 66-70 | Lesional | R | Temporal | Scalp | R | Temporal | R | Temporal | | No | Y |
| sub-PENN49 | PENN | F | 36-40 | Nonlesional | | | Scalp | L | Temporal | L | Temporal | | No | |
| sub-PENN50 | PENN | M | 31-35 | Lesional | B | Temporal | Scalp | L | Frontal; Temporal | L | Temporal | | No | |
| sub-PENN51 | PENN | F | 36-40 | Nonlesional | | | Intracranial | L | Temporal | L | Temporal | IIB | No | |
| sub-PENN52 | PENN | M | 26-30 | Lesional | L | Temporal | Intracranial | L | Temporal | L | Temporal | IB | No | |
| sub-PENN53 | PENN | M | 36-40 | Nonlesional | | | Scalp | B | Temporal | B | Temporal | | No | |
| sub-PENN54 | PENN | M | 31-35 | Lesional | R | Temporal | Scalp | R | Temporal | R | Temporal | | No | |
| sub-PENN55 | PENN | F | 66-70 | Nonlesional | | | Scalp | B | Temporal | B | Temporal | | No | |
| sub-PENN56 | PENN | M | 16-20 | Lesional | L | Temporal | Scalp | L | Temporal | L | Temporal | | No | |
| sub-PENN57 | PENN | F | 41-45 | Nonlesional | | | Scalp | L | Temporal | L | Temporal | | No | |
| sub-PENN58 | PENN | F | 36-40 | Nonlesional | | | Intracranial | L | Temporal | L | Temporal | | No | |
| sub-PENN59 | PENN | M | 26-30 | Nonlesional | | | Intracranial | L | Temporal | L | Temporal | IIIA | No | |
| sub-PENN60 | PENN | M | 36-40 | Nonlesional | | | Intracranial | B | Temporal | B | Temporal | | No | |
| sub-PENN61 | PENN | F | 46-50 | Nonlesional | | | Intracranial | L | Temporal | L | Temporal | | No | |
| sub-PENN62 | PENN | M | 21-25 | Lesional | B | Temporal | Scalp | B | Temporal | B | Temporal | IVB | RNS | Y |
| sub-PENN63 | PENN | F | 26-30 | Lesional | L | Temporal | Scalp | B | Temporal | B | Temporal | | No | Y |
| sub-PENN64 | PENN | M | 31-35 | Lesional | L | Temporal | Scalp | L | Temporal | L | Temporal | IB | No | |
| sub-PENN65 | PENN | M | 51-55 | Lesional | L | Temporal | Scalp | L | Temporal | L | Temporal | IA | DBS | Y |
| sub-PENN66 | PENN | F | 46-50 | Lesional | R | Frontal | Scalp | B | Frontal; Temporal | B | Temporal | | No | Y |
| sub-PENN67 | PENN | F | 46-50 | Lesional | R | Temporal | Scalp | R | Temporal | R | Temporal | | No | Y |
| sub-PENN68 | PENN | M | 36-40 | Lesional | R | Temporal | Scalp | R | Temporal | R | Temporal | | No | |
| sub-PENN69 | PENN | M | 31-35 | Lesional | R | Temporal | Scalp | R | Temporal | R | Temporal | | No | Y |
| sub-PENN70 | PENN | F | 56-60 | Lesional | R | Temporal | Scalp | R | Temporal | R | Temporal | IA | No | |
| sub-PENN71 | PENN | M | 31-35 | Nonlesional | | | Scalp | R | Temporal | R | Temporal | | No | |
| sub-PENN72 | PENN | F | 21-25 | Lesional | L | Temporal | Scalp | R | Temporal | R | Temporal | | No | Y |
| sub-PENN73 | PENN | F | 26-30 | Lesional | L | Temporal | Scalp | L | Frontal; Temporal | L | Temporal | | No | Y |
| sub-PENN74 | PENN | F | 36-40 | Lesional | R | Temporal | Intracranial | R | Temporal | R | Temporal | IIB | No | |
| sub-PENN75 | PENN | M | 46-50 | Lesional | R | Temporal | Scalp | R | Temporal | R | Temporal | | No | Y |

| | | | | | | | | | | | | | | |
|---|---|---|---|---|---|---|---|---|---|---|---|---|---|---|
| sub-PENN76 | PENN | M | 21-25 | Lesional | L | Temporal | Scalp | L | Temporal | L | Temporal | | No | Y |
| sub-PENN77 | PENN | F | 46-50 | Lesional | R | Temporal | Scalp | R | Frontal; Temporal | R | Temporal | | No | Y |
| sub-UOL01 | UOL | F | 61-65 | Nonlesional | | | | | | L | Temporal | | No | |
| sub-UOL02 | UOL | F | 21-25 | Nonlesional | | | | | | L | Temporal | | No | |
| sub-UOL03 | UOL | F | 41-45 | Nonlesional | | | | | | R | Temporal | | No | |
| sub-UOL04 | UOL | F | 51-55 | Lesional | R | Temporal | | | | R | Temporal | | No | Y |
| sub-UOL05 | UOL | F | 36-40 | Lesional | R | Temporal | | | | R | Temporal | | No | Y |
| sub-UOL06 | UOL | F | 36-40 | Lesional | L | Temporal | | | | L | Temporal | | No | Y |
| sub-UOL07 | UOL | F | 16-20 | Nonlesional | | | | | | L | Temporal | | No | |
| sub-UOL08 | UOL | F | 31-35 | Nonlesional | | | | | | R | Temporal | | No | |
| sub-UOL09 | UOL | F | 41-45 | Lesional | L | Temporal | | | | L | Temporal | | No | Y |
| sub-UOL10 | UOL | M | 16-20 | Nonlesional | | | | | | R | Temporal | | No | |
| sub-UOL11 | UOL | F | 21-25 | Lesional | R | Temporal | | | | R | Temporal | | No | Y |
| sub-UOL12 | UOL | F | 36-40 | Lesional | L | Temporal | | | | L | Temporal | | No | |
| sub-UOL13 | UOL | M | 36-40 | Nonlesional | | | | | | L | Temporal | | No | |
| sub-UOL14 | UOL | M | 26-30 | Nonlesional | | | | | | L | Temporal | | No | |
| sub-UOL15 | UOL | F | 36-40 | Lesional | L | Temporal | | | | L | Temporal | | No | Y |
| sub-UOL16 | UOL | F | 16-20 | Nonlesional | | | | | | L | Temporal | | No | |
| sub-UOL17 | UOL | F | 16-20 | Nonlesional | | | | | | L | Temporal | | No | |

### Supplementary Figures

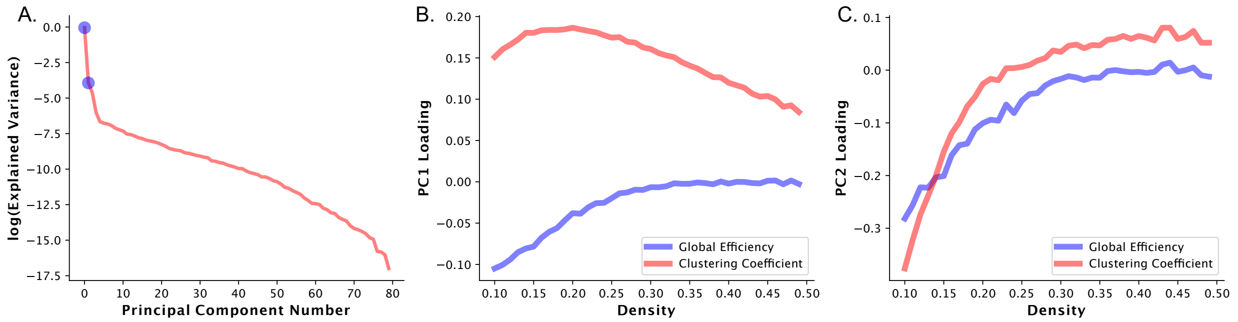


**Supplementary Figure 1 – Principal Components of Integration Segregation Axis:** A. Logarithm of the explained variance as a function of the principal component number. Blue circles represent the values for principal component 1 (PC1) and principal component 2 (PC2). Loadings of B. principal component 1 and C. principal component 2 for the density dependent curves of global efficiency and average clustering coefficient. We can see from B. that PC1 is approximately the tradeoff between global efficiency and average clustering coefficient, particularly at lower densities. From C. we see that PC2 is a weighting of densities for both metrics, where more negative values would suggest increased importance of smaller densities.

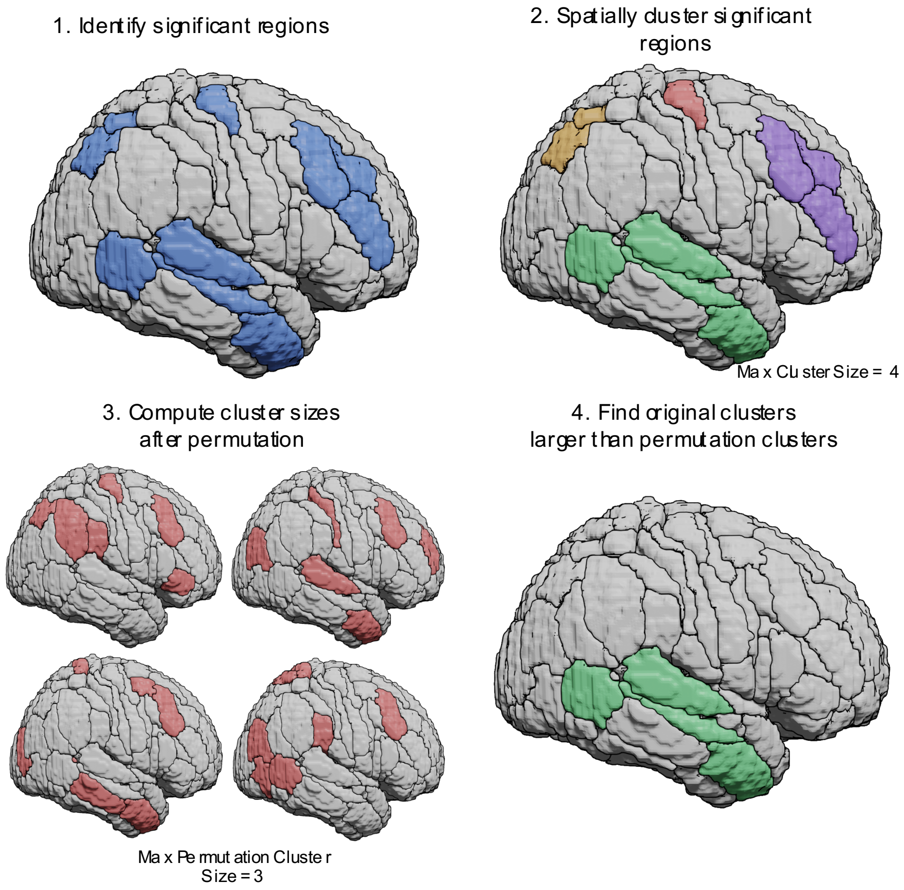


**Supplementary Figure 2 - Diagrammatic explanation of the atlas based suprathreshold test for significance used in this study:** First significant regions are identified between the two groups using a two sample t-test. Then adjacent ROIs that are significant are assigned to the same cluster, and the size of the largest cluster is recorded. In the third step, class labels are permuted and the first two steps are repeated, generating a distribution of maximum cluster sizes. Finally, using the distribution of cluster sizes from the third step, original clusters that are larger than the desired significance level (i.e. larger than 95% of the clusters identified for $p<0.05$) are deemed significant.

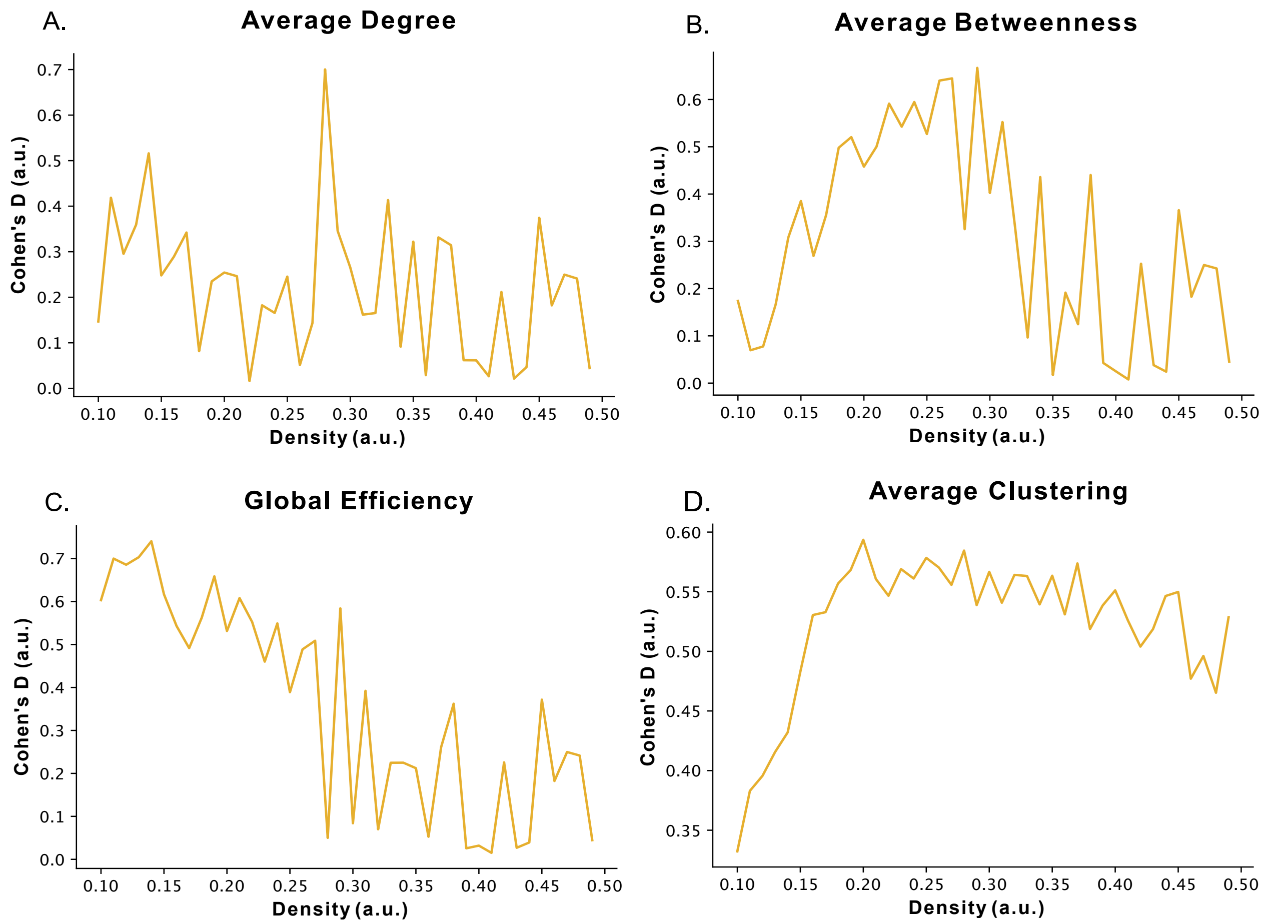


**Supplementary Figure 3 – Cohen's D for network metrics**: Cohen's D across densities between BiTLE and UTLE for whole brain A. average degree, B. average betweenness, C. global efficiency and D. average clustering coefficient.

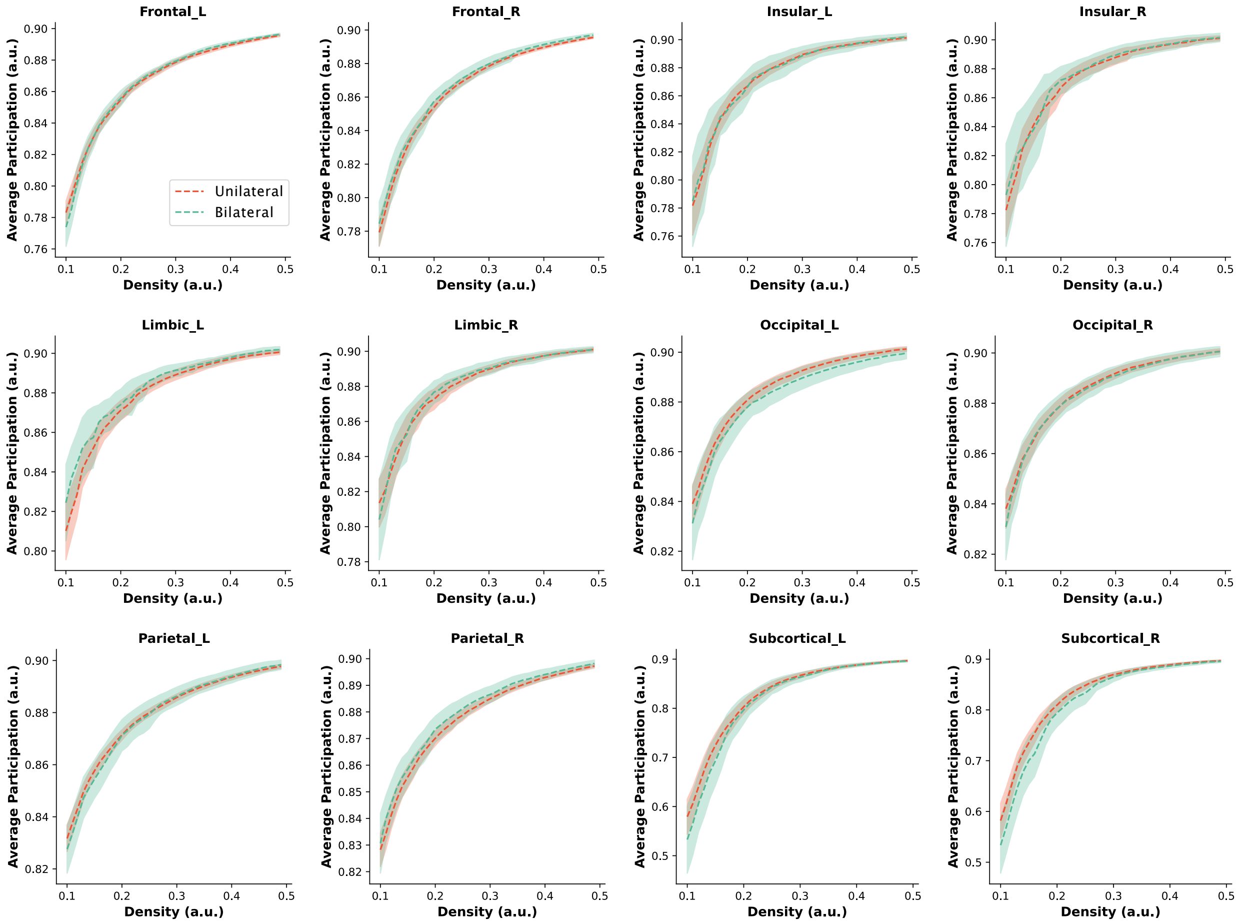


**Supplementary Figure 4 - Participation coefficient across all anatomic communities**. BiTLE (green) and UTLE (red). Dashed lines represent the mean, shaded curve represents 95% CI. * p<0.05.

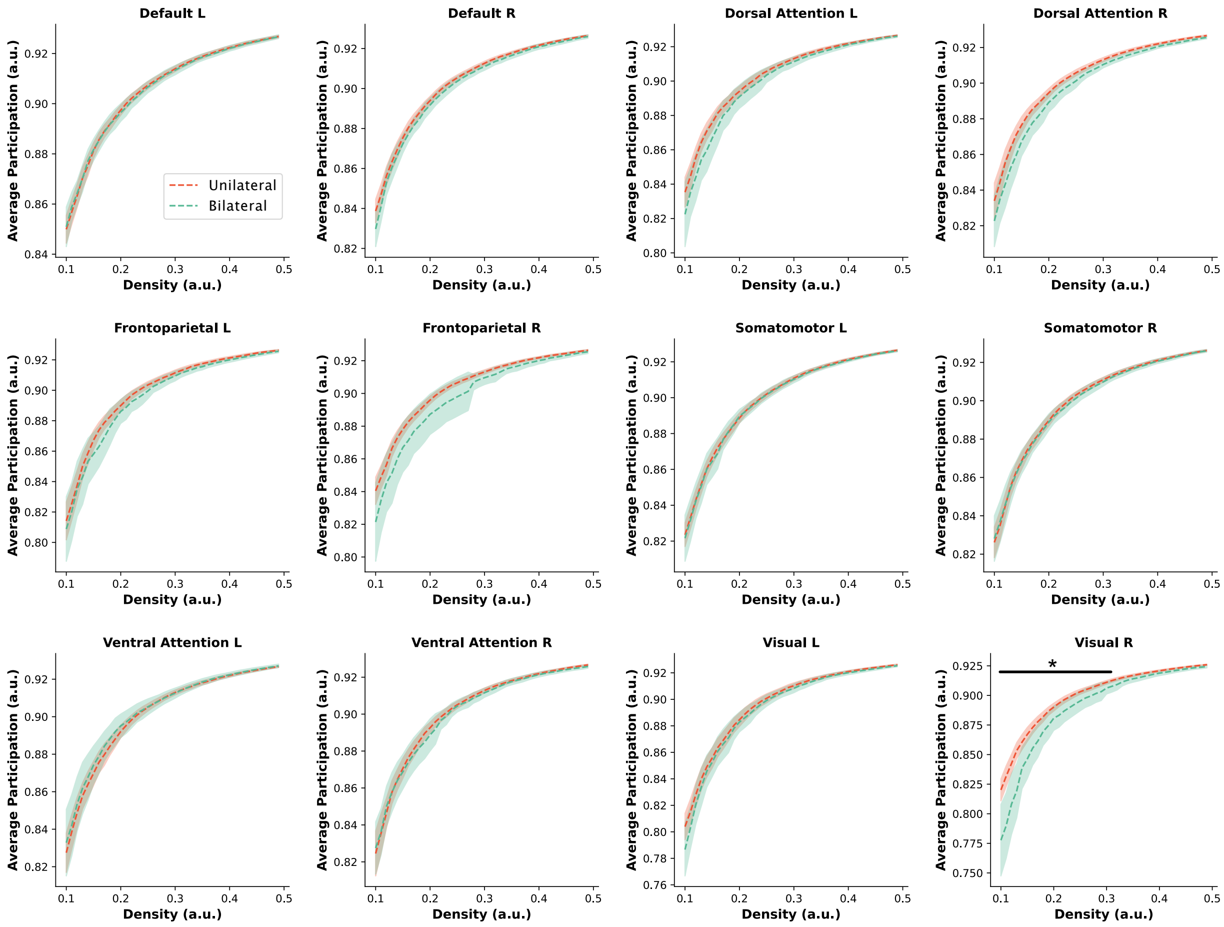


**Supplementary Figure 5 - Participation coefficient across all functional communities.** BiTLE (green) and UTLE (red). Dashed lines represent the mean, shaded curve represents 95% CI. * p<0.05.

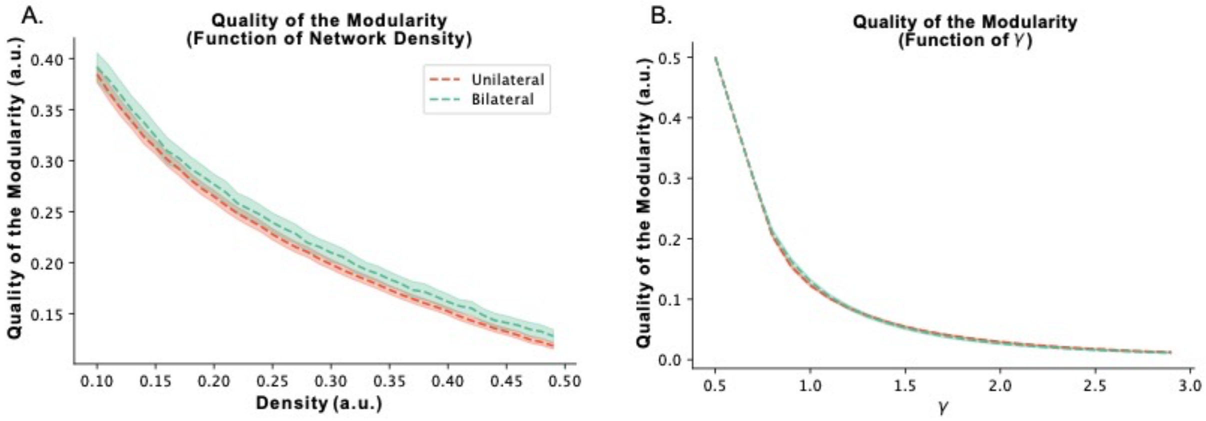


**Supplementary Figure 6** – **Modularity Function Quality:** Quality of the modularity (Q function) as a function of A. network density (significantly different at density values of 0.30-0.40) and B. gamma. BiTLE (green) and UTLE (red). Dashed lines represent the mean, shaded curve represents 95% CI. * $p<0.05$
